# Polygenic prediction of phenotypes with a neural empirical Bayes approach

**DOI:** 10.1101/2025.07.21.25331918

**Authors:** Joshua Weinstock, April Kim, Alexis Battle

## Abstract

Polygenic risk scores (PRS) estimate the expected value of a phenotype based on individual genotypes. Although statistical approaches for calculating PRS have advanced considerably in recent years, few methods incorporate recently generated functional genomics atlases to improve SNP weight estimation. Here, we introduce PRS with a Functional Neural Network (PRSFNN) - a novel approach which uses a neural network in an empirical Bayesian framework to learn the links between SNP functional annotations and SNP weights. By learning these links with a neural network, PRSFNN is able to learn complex, non-linear functions of annotations with minimal assumptions. After curating extensive annotations, including ancestry-stratified allele frequencies, chromatin accessibility across hundreds of developmental and adult cell types, transcription factor binding from ENCODE4, quantitative trait loci, and sequence conservation from Zoonomia, we evaluated PRSFNN on 18 continuous complex traits in the UK Biobank. After benchmarking against other leading PRS methods in an out-of-sample test set, we find that PRSFNN outperforms other PRS methods on 17 of 18 traits. Finally, we show that a low-density lipoprotein PRS estimated with PRSFNN outperforms other PRS methods in the prediction of incident cardiovascular disease. Overall, PRSFNN uses a curated SNP annotation atlas within a neural empirical Bayesian framework to achieve state-of-the-art performance, advancing our ability to predict phenotypic variation from genetic variation.

## 1 Introduction

Understanding how variation in genetic sequence manifests itself in phenotypic variation is a key challenge of human genetics. Polygenic risk scores (PRS), which are aggregations of risk alleles for a given phenotype, offer a genome-wide quantification of the phenotype one would expect given the alleles present for an individual at common variants. These scores typically aggregate the effects from many single-nucleotide polymorphisms (SNPs) into a single individual level estimate of germline predisposition for a disease phenotype. Due to the potential translational and clinical impact of these scores, many statistical methods for estimating the PRS weights have been developed. PRS weights improve upon GWAS weights by applying statistical shrinkage to mitigate overfitting stemming from finite sample sizes. These approaches include pruning and thresholding (P+T) [15], Bayesian approaches with various priors [28, 44, 50, 55], and penalized regression methods [40].

A subset of these methods integrate genomic and regulatory annotations to improve the estimation of the weight of the SNP, which is done by typically pooling information across SNPs that share a given set of annotations. For example, LDpred-funct [41] uses the LDscore baseline model v2.1 [23, 26], which includes regulatory features describing histone marks, enhancer regions, and conservation metrics. AnnoPred [29] uses a similar approach to the variance term of a gaussian prior conditional on functional annotations. Despite the innovations in using functional annotations, existing methods all assume a linear specification of the prior conditional on the annotations. Although linear additive models have proven quite robust in estimating genetic effects on phenotypes, the same observations need not hold when estimating the association between functional annotations and SNP weights.

In this work, we present the Polygenic Risk Score Functionally informed Neural Network (PRSFNN) for polygenic prediction. In contrast to previous efforts, PRSFNN uses a neural network (NN) to learn the functional data informed prior. The NN enables PRSFNN to learn non-linear relationships between the functional annotations and components of the prior. Due to the non-parametric specification of this link, PRSFNN is able to learn complex relationships from heterogeneous collections of SNP annotations, including those that catalog whether a SNP resides in accessible chromatin across cell types [21, 59], AlphaMissense estimates of variant pathogenicity [13], and SNP allele frequencies across genetic ancestries [7]. We demonstrate that PRSFNN outperforms other commonly used PRS methods for the majority of assessed complex traits. In an example of downstream utility, PRSFNN also outperforms other PRS methods when used to predict incident cardiovascular disease. PRSFNN is available today with a scalable implementation in Julia.

## 2 Results

### 2.1 PRSFNN

PRSFNN is an empirical Bayesian method that uses genome-wide association study (GWAS) summary statistics, estimates of linkage disequilibrium (LD), and a database of SNP annotations as input; it does not require individual-level data and can be trained on any population with GWAS summary statistics and a suitable LD reference panel (1). After training the model, PRSFNN estimates the posterior mean of the SNP weights - these weights are then used to compute the final PRS estimate. Prior to training, the SNP weight prior assumes a mixture of two gaussians of mean zero - one with a larger variance than the other. During training, one gaussian will have a fixed variance, and the other gaussian has an adaptive variance based on SNP annotations. After training, the mixture parameter and the variance of the adaptive gaussian are estimated as a function of the SNP weights; this function is learned with a NN that jointly models the mixture parameter and variance term.

PRSFNN is distinguished from other PRS methods that incorporate functional annotations by the use of a NN during the functional prior training process, and by the curation of an extensive set of functional annotations that have been recently generated by large-scale consortia, including ancestry stratified allele frequencies from deeply sequenced genomes [7], cis-regulatory elements from hundreds of fetal and adult cell types [59], transcription factor binding [21], sequence conservation [16], AlphaMissense [13], and QTL annotations [3] (1 A,B, Methods).

In contrast to other Bayesian PRS methods that lack a closed-form analytic posterior [28], we develop an optimized inference scheme using coordinate ascent variational inference (CAVI) [8, 10]. We adapted a previously derived CAVI algorithm for sparse Bayesian linear regression on individual level data to PRSFNN, which included three modifications: 1) exchanging the “spike” component of the “spike-and-slab” prior for a gaussian with small but non-zero variance; 2) exchanging the fixed mixture parameter and slab variance for estimates from the NN; 3) computing the updates of the algorithm using summary statistics instead of individual level data.

### 2.2 Benchmarking polygenic risk score methods

We sought to evaluate the performance of PRSFNN in comparison to other leading PRS methods using a set of samples held out from training. We first trained LDpred2-inf [44], LDpred-funct [41], PRS-CS [28], SBayesRC [61], and PRSFNN on 18 continuous biomarker GWAS performed on British European (EUR) genetic ancestry samples that had previously been generated from UK Biobank (UKB) samples (average *N* = 373*K*). We then used the weights from each trained method to generate phenotype predictions in 27,878 individuals of non-British European ancestry, and assessed performance by comparing predicted phenotypes to the observed phenotypes (2 A).

We compared prediction performance (*R*^2^) for five main methods, finding that PRSFNN yielded the highest prediction accuracy in 17 out of 18 traits (2 B). Notably, PRSFNN substantially outperformed LDpred-funct and SBayesRC, the two methods that incorporate functional annotations, demonstrating the value of its neural network-based modeling. Compared to these annotation-informed methods, PRSFNN achieved a 30.5% average relative increase in prediction performance in the 17 best-performing traits, underscoring its ability to leverage functional data more effectively. The improved performance over SBayesRC, in particular, may be in part attributed to PRSFNN’s robustness to LD reference panels constructed from small sample sizes. SBayesRC uses low-rank LD estimates derived from UKB data; we observed that performance was very sensitive to choice of European ancestry reference panel [61] (A). In contrast, PRSFNN maintained stable performance even when using smaller reference panels such as the 1kGP dataset constructed from 633 individuals of EUR ancestry. Compared to methods restricted to HapMap3 SNPs [33] (LDpred2-inf and PRS-CS), PRSFNN’s use of 6.5 M SNPs, in combination with functional training, led to a 24.4% average relative increase in prediction accuracy across the 17 best-performing traits.

**Fig. 1.**
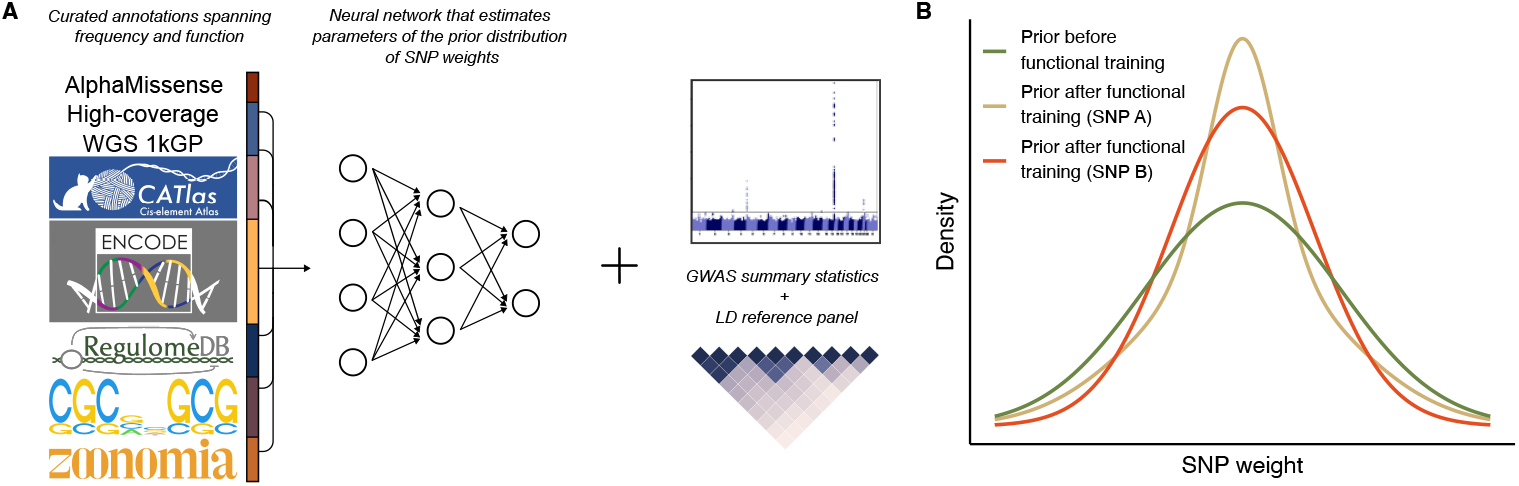
Polygenic risk scores with a functional neural network (PRSFNN). A) Architecture of the functional NN prior. B) On exemplar SNPs, the prior densities are included for: 1. the prior without training; 2. The prior after functional tuning for SNP A. ; 3. The prior after functional tuning for SNP B.

The average CPU hours and user run time for all five methods are reported in A. We report only the time required to estimate posterior mean causal effect sizes, excluding the time to compute LD matrices, as each method handles LD construction differently.

### 2.3 The Effects of Functional Training on PRS performance

To estimate the relative contribution of functional training to PRS performance, we then systematically compared PRSFNN with and without functional training across 18 phenotypes. To assess the performance of PRSFNN without functional training, we used a mixture of two gaussians as the prior without incorporating the NN; all other algorithmic properties were unchanged (Methods). We observed that changes in performance were highly heterogeneous: lipid phenotypes improved by an average Δ*R*^2^ = 0.013 (5.3% relative gain) with functional training over the naive prior, while blood cell phenotypes had a larger gain in performance (Δ*R*^2^ = 0.033, 19.5% relative) (3 A). In contrast, anthropometric phenotypes showed only a minimal average improvement in bone mineral density, BMI and systolic blood pressure (Δ*R*^2^ = 0.00092). Using orthogonal heritability estimates for 15 out of 18 traits [48], we observed that lower heritability traits benefited more from functional training (3 B), whereas the most heritable traits showed smaller increases in performance, which may reflect the absence of relevant functional annotations for causal tissues in these phenotypes.

**Fig. 2.**
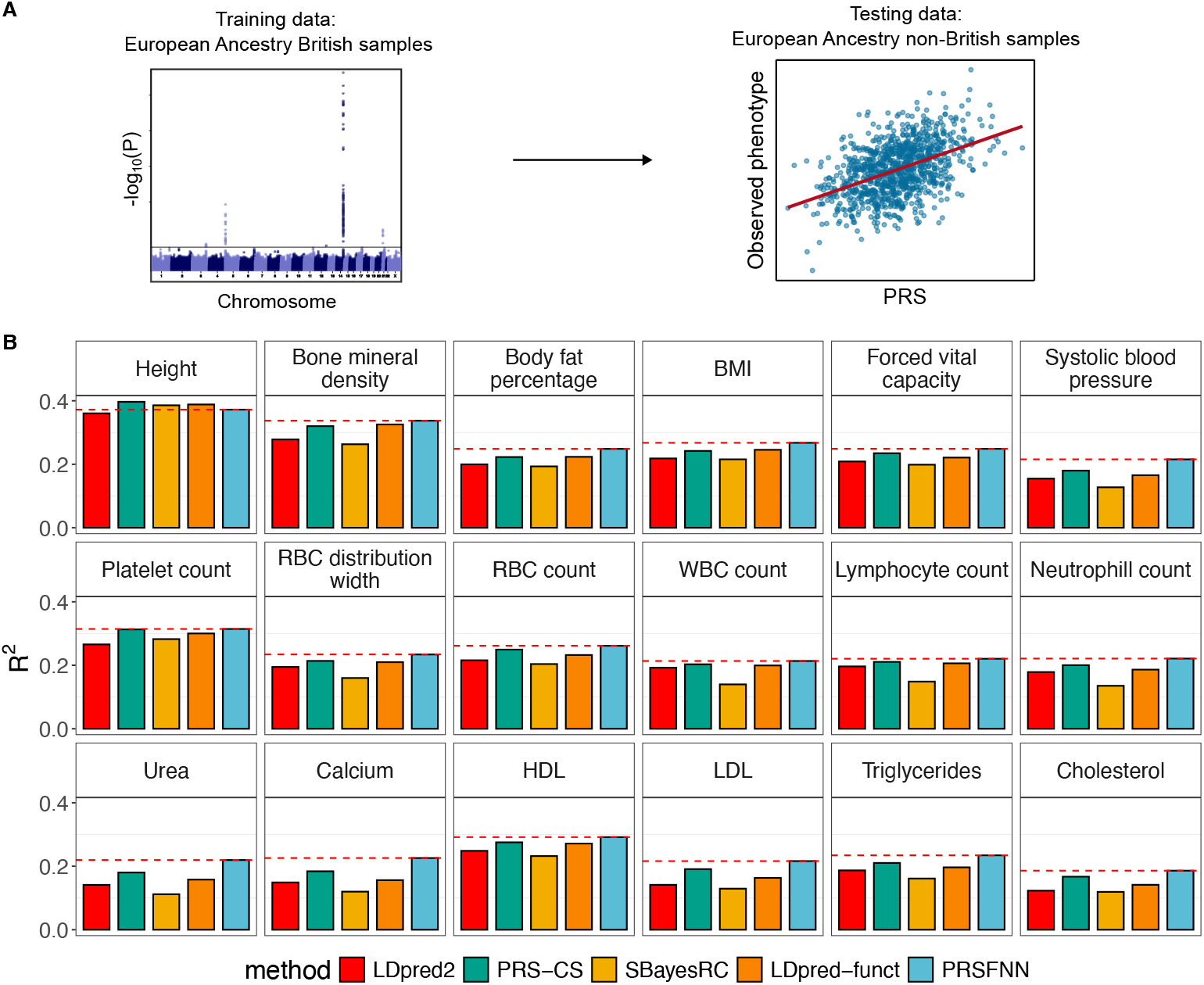
Accuracy of 5 PRS methods across 18 UK Biobank quantitative traits. A) Evaluation was performed using British ancestry individuals for training, and predictive performance was assessed on non-British white individuals. PRSs were computed using genotypes from the test set, and prediction accuracy was measured using R^2^. B) We report predictive accuracy for LDpred2-inf, PRS-CS, SBayesRC, LDpred-funct and PRSFNN. The red dashed line indicates the performance of PRSFNN. Abbreviations: BMI (body mass index), RBC distribution width (red blood cell distribution width), RBC count (red blood cell count), WBC count (white blood cell count), HDL (high-density lipoprotein), LDL (low-density lipoprotein).

We next performed an enrichment analysis to examine which features were associated with SNPs prioritization by the functional prior (Methods). SNPs favored by the NN were defined as those with a high ratio of the two posterior point–estimates, the adaptive gaussian component and the fixed component; the remaining SNPs were considered unprioritized by the NN. For each phenotype, we fitted a Firth’s logistic regression model predicting this binary outcome from the full set of annotations; this model provides a simplified, surrogate model of the NN that we trained to facilitate interpretation. To capture non-linear effects, AlphaMissense pathogenicity and the number of accessible cell types were modeled using linear splines that model step functions (Methods). Across all traits, rare variants (minor-allele frequency *<* 5%) in regulatory elements with moderate, multi-cell-type chromatin accessibility (accessible in greater than 10 cell types, but fewer than 50) were significantly enriched among the prioritized SNPs, whereas common SNPs were depleted (3 C). In particular, SNPs accessible in 10-50 cell types showed greater enrichment than those accessible in over 50 cell types, underscoring that cell-type specificity rather than broad ubiquity drives functional prioritization. Moreover, AlphaMissense pathogenicity scores exhibited a non-linear relationship – SNPs with non-zero, but ‘benign’ (¡ 0.34) scores were prioritized, in contrast to variants of uncertain classification (AlphaMissense scores from 0.34 to 0.56), which were depleted. In summary, the non-linear effects of variant pathogenicity and chromatin accessibility underscore the value of a non-parametric link between SNP annotations and parameters of the prior distribution.

**Fig. 3.**
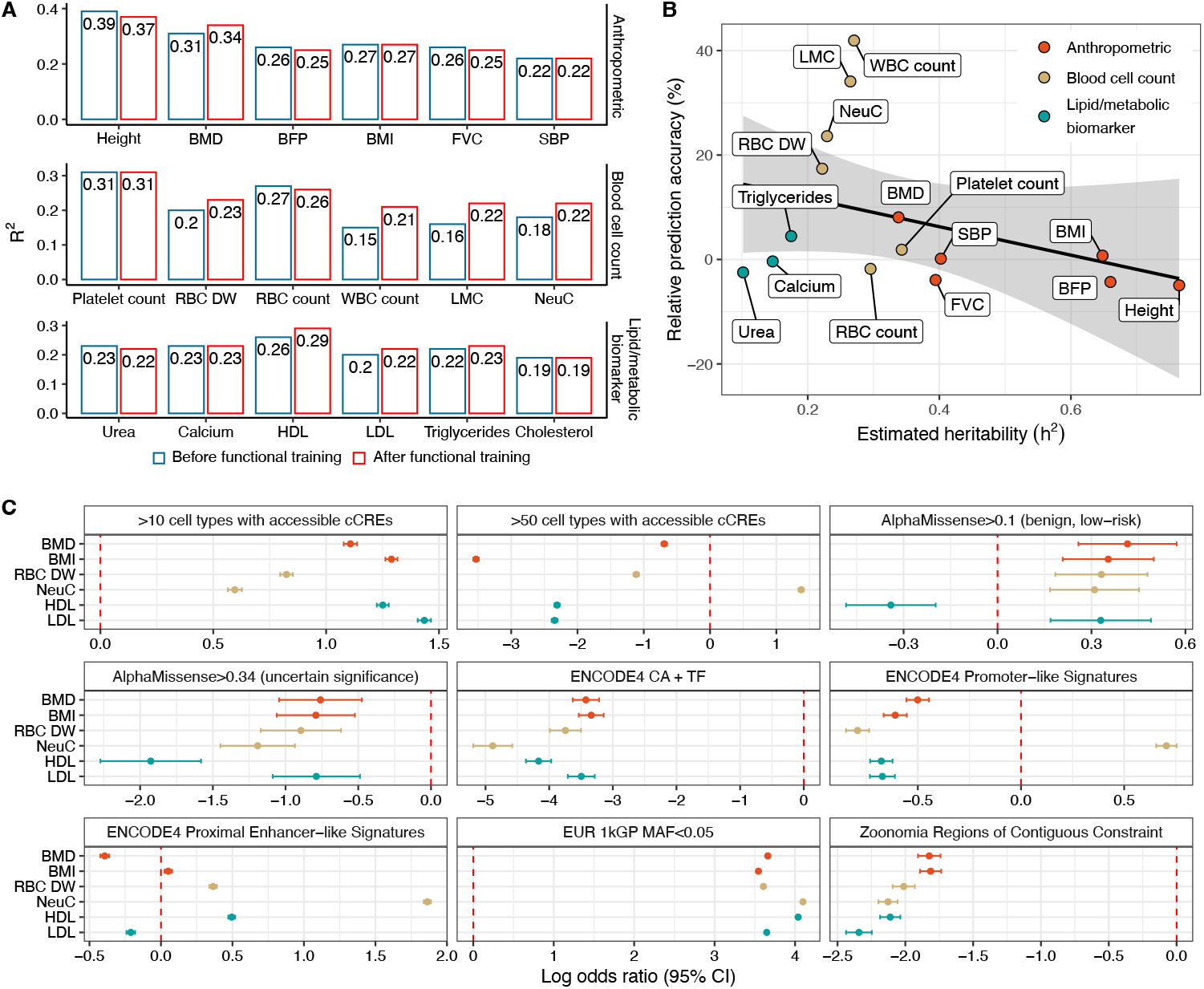
Functional training prioritizes rare variants in accessible regulatory elements. A) PRSFNN with functional training (red), on average, is superior to without incorporation of the NN (blue), as measured by R^2^. Phenotypes are grouped into three categories: anthropometric, blood cell counts and biomarkers. Abbreviations: BMD (bone mineral density), BFP (body fat percentage), BMI (body mass index), FVC (forced vital capacity), SBP (systolic blood pressure), RBC DW (red blood cell distribution width), RBC count (red blood cell count), WBC count (white blood cell count), LMC (lymphocyte count), NeuC (neutrophil count), HDL (high-density lipoprotein), LDL (low-density lipoprotein). B) Traits with relative lower heritabilities tend to benefit more from functional training. C) Logistic regression was used to identify features enriched among the SNPs most strongly favored by the NN (top 1% by the ratio of the two posterior point–estimates for each SNP). The binary outcome was membership in this top percentile and predictors were: AlphaMissense pathogenicity score, EUR allele frequency (1kGP), number of cell types with accessible chromatin (cCRE), regulatory signatures from ENCODE4 and RegulomeDB, TF PWM motif hits, and regions of contiguous constraint (Zoonomia). A subset of the most informative predictors is shown; full model and results are detailed in Methods and C.

### 2.4 LDL PRS informs CAD risk stratification

**Fig. 4.**
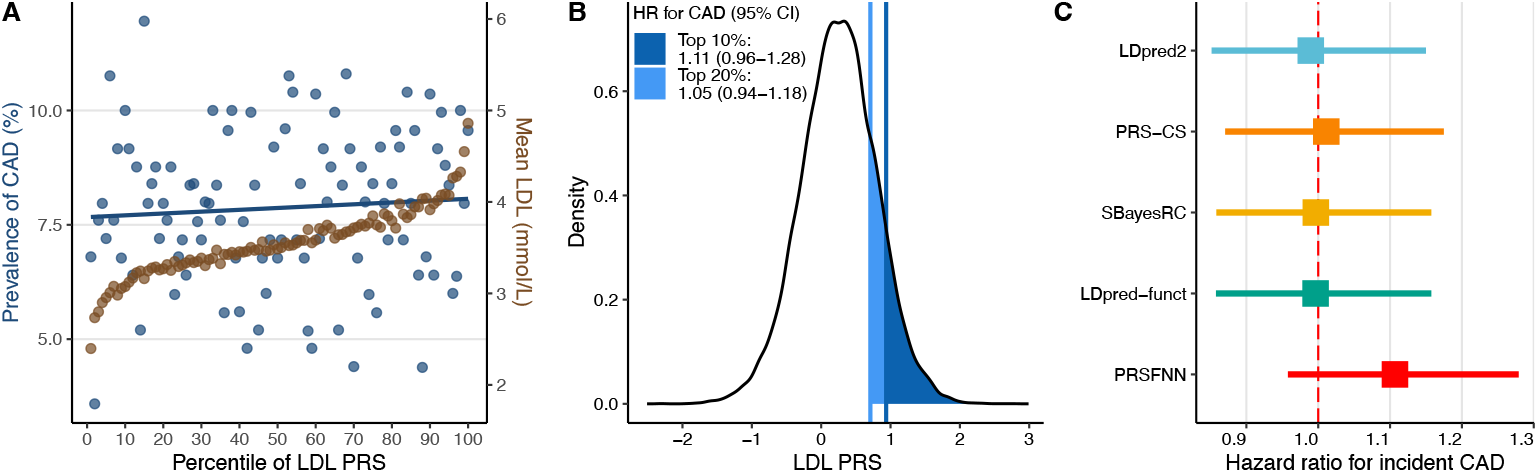
Stratification by PRSFNN LDL PRS provides the most predictive utility for CAD risk. A) Blue dots indicate the prevalence of CAD across percentiles of the genome-wide PRS for LDL in the UK Biobank testing cohort. Brown dots represent the average LDL cholesterol levels within each corresponding PRS percentile. B) Individuals in the top 10% and 20% of the LDL PRS distribution had a higher risk of CAD compared to the remaining 90% and 80% of the population, respectively, as measured by hazard ratios. C) Cox proportional hazards models were used to assess the association between LDL PRS and time to CAD events, comparing individuals in the top decile (high LDL PRS) to those in the bottom 90%. Each model was adjusted for age at assessment, sex, BMI, smoking status, diabetes status, statin use, systolic and diastolic blood pressure, and the top 10 genetic principal components. Hazard ratios together with their 95% confidence interval are shown for each of the five PRS methods.

We then conducted a survival analysis to assess the potential clinical utility of LDL PRS for coronary artery disease (CAD). Specifically, we examined whether individuals with higher genetically predicted LDL cholesterol levels, estimated using PRS from five different methods, experience an earlier onset of CAD events. In our models, incident CAD events were treated as the event phenotype, and survival time was defined from the date of UKB enrollment to the earliest of CAD diagnosis, death, or censoring. CAD cases and controls were defined from ICD10 codes the “phecode” system (phecode 411, “Ischemic heart diseases”, Methods). Individuals without a CAD diagnosis were right-censored at the time of last follow up or death.

In the UKB test set, which included 26,909 individuals and 2,138 CAD cases, LDL PRS percentiles were strongly associated with average LDL cholesterol levels (4 A). Similarly, higher LDL PRS was associated with increased CAD prevalence across percentiles. To evaluate stratification based on PRS, we defined a dichotomized PRS variable comparing individuals in the top decile to the remaining 90% of the cohort. Individuals in the top 10% of the LDL PRS distribution had a hazard ratio (HR) (95% CI) of 1.11 (0.96-1.28) for CAD, and those in the top 20% had a HR of 1.05 (0.94-1.18), relative to the remaining population (4 B).

Among all methods, PRSFNN showed the strongest risk stratification, with individuals in the top decile exhibiting a significantly increased risk of CAD after adjusting for age at assessment, sex, BMI, smoking status, diabetes status, statin use, systolic and diastolic blood pressure, and the top 10 principal components (4 C). Overall, these results indicate that improved PRS performance with PRSFNN is likely to translate to improved disease risk stratification.

## 3 Discussion

Although human genetics has progressed to an era where population-scale sequencing is routinely performed [1, 6, 52, 53], phenotype prediction from genotype remains challenging. In this work, we integrate a parametric Bayesian PRS approach with a neural network, PRSFNN, to leverage large-scale functional data that have been produced by consortia in recent years [3, 7, 13, 16, 21, 47, 59]. After comprehensive benchmarking of PRSFNN with other leading PRS methods, we observed that PRSFNN most accurately recapitulated continuous biomarker phenotypes in an out-of-sample test set of individuals. We observed that this can also support utility in translational contexts; when evaluating LDL PRS estimates as predictors in a survival analysis of incident cardiovascular disease, we found that PRSFNN had the largest effect size. This implies that if individual groups were stratified based on polygenic predisposition to hyperlipidemia – an early screening procedure which may become routine practice in the coming decades [5, 39, 54] – PRSFNN would provide the most useful estimates among those benchmarked.

We emphasize that although our approach is not the first to incorporate functional annotations [11, 30, 41, 61], it provides the first implementation with a completely agnostic link between functional annotations and the parameters of the prior distribution. This enables PRSFNN to learn potentially complex interactions between distinct classes of annotations (e.g., allele frequencies and chromatin accessibility). This is likely to be especially important for phenotypes where a Bayesian prior is most useful: those collected from cohorts with limited sample sizes or those with limited heritability dispersed over a very large number of causal variants. In these cases, the GWAS data are in a sense less informative, lending more weight to the prior. This suggests that until human genetics progresses to a more equitable state where large-scale biobanks are less biased towards European genetic ancestry ascertainments, functionally informed Bayesian PRS are likely to be especially useful in smaller cohorts with more diverse sampling strategies.

We note that our results contrast with a recent report that describes the lack of utility in incorporating deep learning (DL) methods into PRS [34]. In this effort, a PRS was constructed by using a NN to estimate the expected value of the phenotype by using a NN as the estimation method. We show that the careful integration of traditional, Bayesian PRS formulation, where the phenotypic risk is still assumed to be a linear function of genotype, with a DL component used as a prior on weights shows highly promising results for phenotype prediction. This observation highlights that wholesale claims regarding the utiltity of DL in PRS are likely to be somewhat misleading; DL most importantly represents a non-parametric estimator of some quantity, and traditional PRS algorithms have several components where a non-parametric addition stands to add value, such as in our priors. Similarly, it may be useful to specify the residual variance term as a function of a neutral network. DL is likely to be especially useful when the true functional form departs from linearity by a significant degree and when manual feature engineering is likely to miss important combinations of features.

We remark that there is no particular reason from a statistical learning perspective as to why advances are most likely to emerge from future iteration with NNs; although we found success with the implemented architecture, we are not aware of any reason why a similar non-parametric estimator (e.g., gradient boosting or Bayesian additive regression trees [12, 14, 19, 25]) would not yield comparable, or perhaps even superior, results. However, one particular advantage of DL is the recent emergence of an incredible breadth of DL related tooling, which facilitates development of scalable DL methods [4, 31, 32, 43].

To interpret the functional prior, we used a surrogate logistic regression model to identify SNP annotations that were enriched among those SNPs that were highly prioritized by the NN. Concordant with past reports that partition heritability based on SNP annotations [22, 27] we find that SNPs in tissue-specific regulatory elements are enriched for having larger effect sizes. However, we observed non-linearity with respect to the number of cell types in which a regulatory element is active; we observed that SNPs in regulatory elements that were shared across numerous (¿ 50) cell types were less likely to be prioritized than SNPs in regulatory elements active in 10-50 cell types, which may suggest that ubiquitously active regulatory elements are depleted of large effect variants. This observation is concordant with a recent comparative analysis of GWAS and RVAS hits, which reported that GWAS variants are enriched for trait-specificity [51]. Similarly, we also observed non-linearity with respect to the predicted pathogenicity of missense variants; missense variants that were predicted by AlphaMissense to have modest impact were more likely to be prioritized by the prior than more deleterious variants. Although coding variants are clearly enriched for heritability in GWAS, highly deleterious variants are likely so infrequent as GWAS hits that functional priors are likely to underestimate their importance.

Given the modular nature of our functional prior, further opportunities exist to incorporate recent advances in artificial intelligence (AI) approaches that predict molecular phenotypes from DNA sequence [2, 20, 35, 37, 42, 63]. We anticipate that future iterations of these models, particularly those that show promise in predicting the effects of inter-individual variation, may prove especially powerful as functional annotations for rare variants.

Beyond curating additional sources of functional annotations, we identify five directions for future iterations. First, we anticipate that approaches that construct ensembles over ancestry-specific models will likely yield improved portability of PRS predictions [45, 57, 58]. Second, possibilities for jointly modeling SNPs and indels along with other forms of germline genetic variation, particularly structural variants (those over 50 bp in length). The integration of structural variants within PRSFNN is likely to prove especially useful, given the recent dramatic increase in long-read sequencing and the development of graph based pan-genome efforts [36]. Third, in parallel with recent efforts to improve prediction of phenotype from sequence, estimation of uncertainty should also proceed in concert; conceptually, it is entirely possible to also specify the residual variance term in PRSFNN as a function of a NN; non-constant variance terms (heteroscedasticity) have been previously explored in generalized linear mixed model contexts [17]. Fourth, PRSFNN may benefit from multi-phenotype extensions that enable the model to learn from multiple phenotypes with shared parameters. Finally, in an analogous manner to how genomic foundation models learn representations or embeddings among genes, the hidden units of the functional prior may also provide useful, phenotype-specific representations that provide substrate for subsequent task-specific fine-tuning, e.g., other phenotypes, or gene-based testing.

Overall, PRSFNN provides state-of-the-art performance in predicting phenotypes from sequence using a neural empirical Bayesian framework. PRSFNN is available in an open source, scalable Julia implementation.

## 4 Methods

### 4.1 Likelihood

We model the conditional probability of a phenotype random vector **Y** as follows:

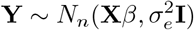

which implies the likelihood:

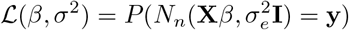

Where **y** is a length *n* vector representing the observed phenotype values, **X** is a *n* x *p* matrix containing centered genotype data, *β* is length *p* random variable representing the weights of SNPs, and 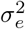 is the residual variance.

#### 4.1.1 PRSFNN

Computing this likelihood requires access to individual level data. We instead use an approximate summary statistic likelihood introduced in [64] that only requires marginal test statistics from a GWAS and estimates of linkage disequilibrium (LD) from a suitable reference panel.

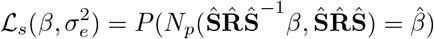

Where 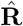 is an estimate of the LD matrix, 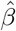 are the genome-wide association study (GWAS) effect estimates, and **Ŝ** are the standard errors of the GWAS summary statistics. We note that the population of the reference panel used to compute 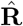 should be the same as the population where the GWAS was performed. In practice, we also first standardize 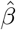 based on the estimated residual variance and allele frequencies.

#### 4.1.2 Estimating residual variance

We estimate the residual variance, *σ*_*e*_, from summary statistics following [56]:

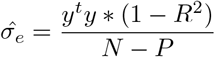

Where 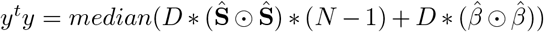, and *D* is defined as the diagonal elements of *X*^*t*^*X*.

#### 4.1.3 LD Computation

We compute LD using genotypes from the high-coverage sequencing of the 1000 Genomes Project [7]. Rather than compute LD across the entire genome, which has compute complexity *O*(*np*^2^), we instead compute LD estimates separately within independent LD blocks. To define independent LD blocks, we used the estimates of recombination rates from [49]. We use a simple ridge-like estimator of the covariance, i.e.,

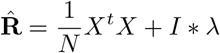

The regularization is needed to ensure that 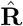 is positive definite. A recent theoretical analysis of block-wise LD estimators indicated that although block-wise estimators perform worse than methods that use the full genotype matrix, the difference is fairly small for phenotypes with *h*^2^ values less than 40% [60].

### 4.2 Prior

Over the SNP weights, we assume the following prior distribution:

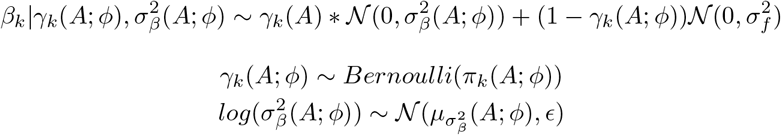

Where **A** is a *p* x *l* matrix of annotations and 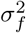 represents a small, non-negative hyper-parameter.

The prior inclusion probability *π*_*k*_(*A*; *ϕ*) and the slab variance 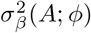 are functions of the matrix of *A*, where *ϕ* denotes the hyper-parameters of these deterministic mappings. The notation *f*(*A*; *ϕ*) is used to emphasize that the mixture parameter and adaptive variance are functions of the annotations *A. ϵ* is a training hyper-parameter that we set to 1.0. The mappings will be computed via a neural network (NN). Thus, the prior conditional on the annotations for the *kth* SNP is mixture of two gaussians; one with a fixed variance representing a polygenic background, and the other with a learned adaptive variance. Due to the use of a NN to compute the mixture parameter and adaptive component variance, the prior distribution has more flexibility than standard Bayesian priors.

This leads to a joint log probability:

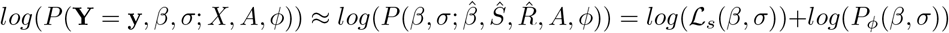

### 4.3 Posterior computation

We take the following surrogate posterior approximation to the true posterior *π*(*β*) :

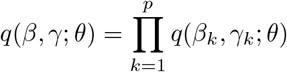

Where

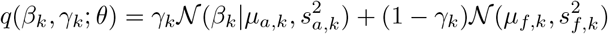

And 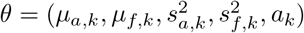 Conditional on an given

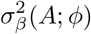

and,

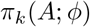

we take the coordinate-ascent variational updates following Carbonetto and Stephens [9] after exchanging ℒ_*i*_ for ℒ_*s*_ and following the changes from their prior.

#### Algorithm 1 Coordinate-ascent variational updates

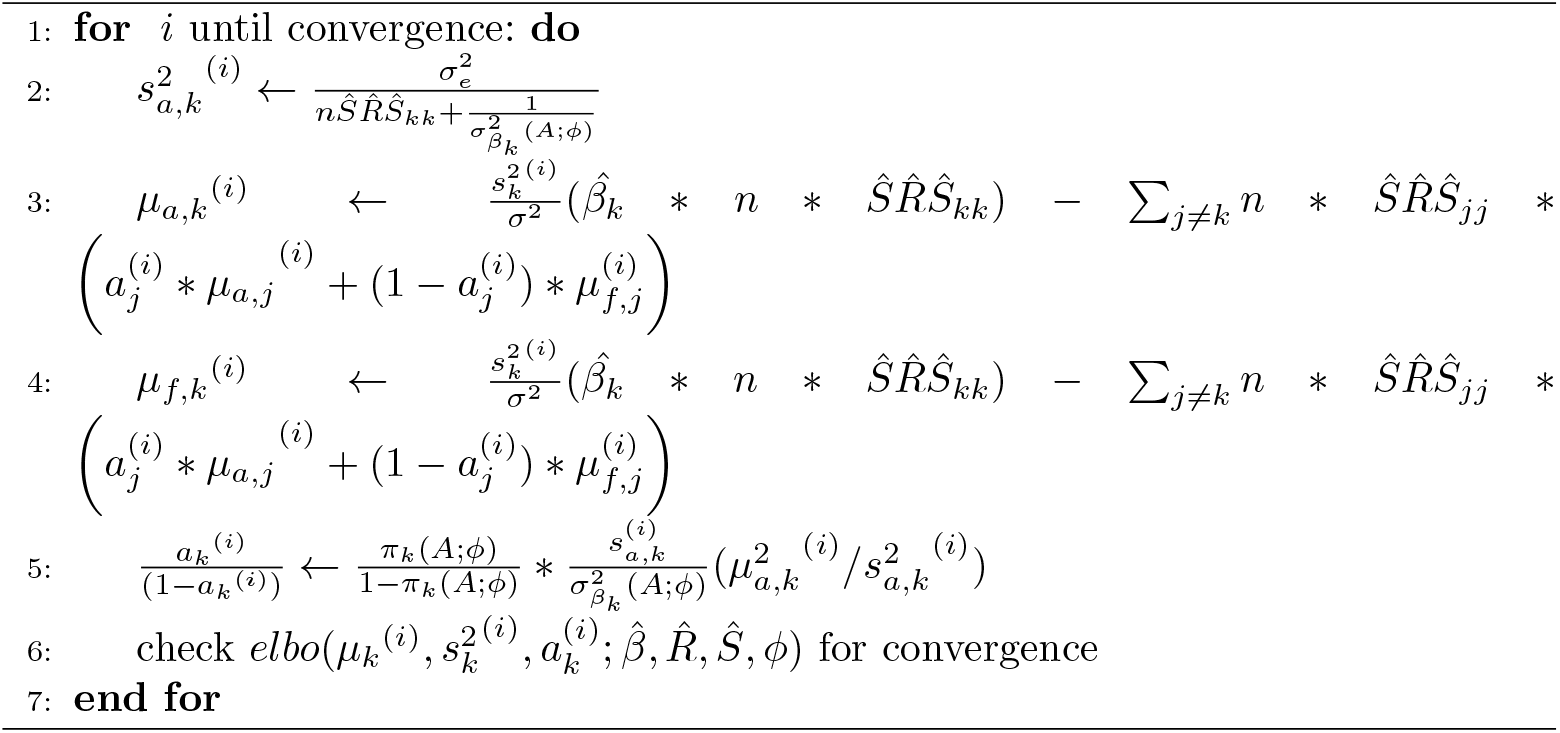

The point estimate most useful for PRS is the expectation of the posterior mean for the SNP coefficient, which is a weighted average of the estimates of the mean for the adaptive and fixed components:

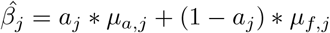

### 4.4 Estimation of hyper-parameters

*π*_*k*_(*A*; *ϕ*) and 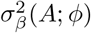 are unknown mappings. We take both mappings to be two output neurons of a single NN parameterized by weights *ϕ*. We optimize *ϕ* within the context of a variational-EM algorithm:

#### Algorithm 2 Variational-EM algorithm

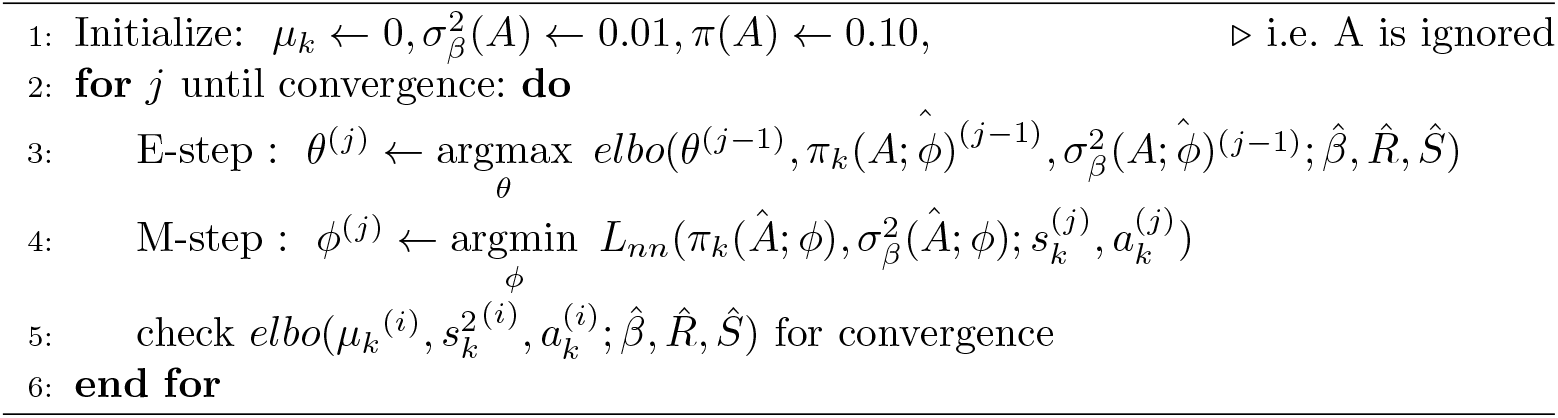

Where 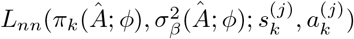 is defined as:

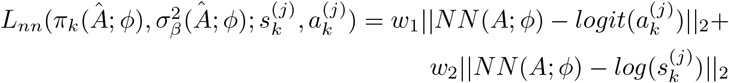

where *w*_1_ and *w*_2_ represent weights for the two components. In practice, we run this for a single iteration.

### 4.5 Neural network architecture

We use an architecture that consists of a single hidden layer with a smooth non-linear activation function (Softplus). Parameters are optimized using the AdamW optimizer [38]. Weights are initialized using Glorot-normal initialization with a small gain (0.005) to promote stable early training. Biases in the final layer are initialized to reflect weak priors: *log*(0.001) for the effect variance component, and *logit*(0.1) for the causal probability.

### 4.6 Block-wise training scheme

The model is trained using a block-wise stochastic optimization scheme. Each training epoch corresponds to a single LD block, from which both the annotation matrix and associated summary statistics are extracted. This approach allows for memory-efficient training on genome-wide data and implicitly leverages LD structure. To promote generalization and mitigate overfitting, we hold out a random subset of LD blocks as a validation set.

### 4.7 Genomic and functional SNP annotations

We included numerous genomic and single cell-type specific regulatory annotations spanning hundreds of cell types from both fetal and adult human tissues.

1. Ancestry stratified minor allele-frequencies [7]
2. Single-cell-type specific chromatin accessibility information from [59]
3. Conservation metrics from Zoonomia [16]
4. Regulatory functions from RegulomeDB [3]
5. Transcription factor binding sites [47]
6. Predictions from AlphaMissense [13]
7. Regulatory element annotations from ENCODE4 [21]

### 4.8 Post-training model interpretation

Logistic regression was used to identify SNP annotations most prioritized by the NN. We used 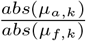 for each SNP as a simple measure to quantify prioritization by the functional prior. Percentile-thresholding was used to label SNPs that were most strongly prioritized by the functional prior. The binary outcome was membership in this top percentile and we included the following predictors: AlphaMissense pathogenicity score, EUR allele frequencies (1kGP), number of cell types with accessible chromatin (cCRE), regulatory signatures (ENCODE4 and RegulomeDB), TF PWM motif hits, and regions of contiguous constraint (Zoonomia). EUR allele frequencies were recoded as common or rare using a 5% minor-allele frequency threshold. Zoonomia ultra-conserved elements were omitted due to its extreme rarity. To capture non-linear effects, we transformed both the AlphaMissense pathogenicity score and the number of accessible cell types into piecewise linear functions using linear splines at biologically meaningful cut-points and included in place of a single linear term. Specifically, AlphaMissense splines were created with pathogenicity score *>* 0.01 (benign, lowest-risk), *>* 0.10 (benign, low-risk), and *>* 0.34 (uncertain significance) and number of accessible cell types splines at accessible cell types *≥*5, *≥*10. *≥*20 and *≥*50. AlphaMissense pathogenicity score *>* 0.56 (pathogenic) was omitted due to rarity.

### 4.9 Survival analysis

To evaluate the relationship between PRS and CAD onset, we applied Cox proportional hazards regression, which estimates the association between time-to-event outcomes and predictors. We studied the incidence of Ischemic heart diseases, with diagnoses based on the International Classification of Diseases (ICD) 10 [18]. Individuals with prevalent disease, defined as having a CAD diagnosis prior to enrollment, were excluded. In the survival analysis, participants were censored at the time of death or at the hospital inpatient data censoring date of October 31, 2022. We reasoned that participants who reported using “cholesterol-lowering medication” were likely taking statins. In summary, among 26,909 individuals, a total of 2,138 incident CAD cases were observed. To assess risk stratification, we included a dichotomized PRS variable comparing individuals in the top decile of PRS to the remaining 90% of the cohort. Each model was adjusted for age at assessment, sex, BMI, smoking status, diabetes status, statin use, systolic and diastolic blood pressure, and the top 10 genetic principal components.

### 4.10 Compared methods

#### 4.10.1 LDpred2-inf

LDpred is a Bayesian method for deriving posterior mean SNP effect sizes from GWAS summary statistics, while accounting for local LD using an external reference panel [44]. LDpred2-inf provides an analytical solution under the infinitesimal model of LDpred where the genetic architecture model assumes all SNPs have non-zero contribution of the phenotype variance. It models SNP effect sizes with a point-normal prior: a fraction *p* of SNPs have non-zero effects drawn from *N* (0, *h*^2^*/Mp*), while the remaining have effect size zero. Given an LD matrix R, GWAS marginal effects 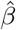, and parameters proportion of causal variants *p* and SNP heritability *h*^2^, LDpred uses Gibbs sampling to estimate the posterior mean effect sizes beta.

#### 4.10.2 LDpred-funct

LDpred-funct extends the LDpred2-inf model by incorporating functional genomic information to refine SNP effect estimates [41]. LDpred-funct integrates trait-specific functional enrichments derived from the baseline-LD model [23, 24], which includes annotations such as coding, conserved, regulatory, and LD-related features. These enrichments are estimated using stratified LD score regression (S-LDSC) [23] applied to the training GWAS summary statistics. SNPs are assigned prior variances proportional to their expected per-SNP heritability 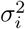, and effect sizes are modeled as 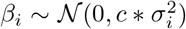 where *c* is a scaling constant ensuring that the total prior variance sums to the SNP-heritability *h*^2^. We used the infinitesimal model version that directly outputs analytically derived posterior mean effect sizes using functional priors without regularization via cross-validation.

#### 4.10.3 PRS-CS

PRS-CS is built under a Bayesian framework that models SNP effect sizes using a continuous shrinkage prior, allowing for marker-specific adaptive shrinkage while accounting for LD [28]. PRS-CS employs a global-local shrinkage prior, where SNP effect sizes are subject to a global scaling parameter *ϕ* and individual marker-specific shrinkage parameters *ψ*_*j*_, which follow gamma distributions *ψ*_*i*_ *∼ Gamma*(*a, δ*_*j*_) and *δ*_*j*_ *∼ Gamma*(*b*, 1). PRS-CS requires a tuning dataset to select the optimal *ϕ* from a pre-defined grid (0.0001 to 1) to maximize prediction accuracy. We used the default parameters *a* = 1 and *b* = 0.5 and fixed *ϕ* = 1*e* − 2 as proposed by the authors.

#### 4.10.4 SBayesRC

SBayesRC is another Bayesian polygenic prediction method that extends SBayesR by integrating functional genomic annotations and enabling the analysis of all imputed common variants genome-wide [62]. Like SBayesR, SBayesRC models SNP effect sizes using a mixture of four zero-mean normal distributions with varying variances, capturing different degrees of effect sparsity. It improves upon this by employing a multicomponent annotation-dependent prior, allowing functional annotations to influence both the probability that a SNP is causal and the distribution of its effect size. SBayesRC utilizes a low-rank approximation of the LD matrix based on eigen-decomposition within quasi-independent LD blocks to efficiently handle millions of SNPs.

### 4.11 Summary statistics and polygenic prediction

UKB GWAS summary statistics for all quantitative traits were accessed from the Neale Lab GWAS v2 release (http://www.nealelab.is/uk-biobank/). We mapped the genetic markers to the Genome Reference Consortium human genome build 38. For LDpred-funct and SBayesRC, we converted the genomic coordinates based variant IDs to dbSNP [47] rs IDs to be compatible with the baseline-LD model identifiers. For LDpred2-inf and PRS-CS, we restricted the genetic markers to the HapMap3 panel as suggested by the authors. We used the 1kGP European sample (*N* = 633) as the external LD reference panel.

All phenotypes were pre-adjusted for age, sex and the first 10 principal components. All methods were used to obtain the posterior mean effect sizes using British ancestry individuals as training data. PRS was calculated and the predictive performance was assessed using genotypes from in an independent testing set of non-British ancestry white individuals. The prediction *R*^2^ was obtained from linear regression of adjusted phenotypes on the PRS.

## Data Availability

Access to the UK Biobank resource is available via application (https://www.ukbiobank.ac.uk/). 1000 Genomes data were obtained from https://www.internationalgenome.org/data/

https://www.internationalgenome.org/data/

## 5 Code Availability

1. PRSFNN Julia module [code](https://github.com/weinstockj/PRS).
2. SNP annotation [code](https://github.com/weinstockj/SNP_annotations).
3. LD Reference panel [code](https://github.com/weinstockj/LD_REFERENCE_PANEL).
4. [Code](https://github.com/weinstockj/process_neale_lab_GWAS) to process GWAS summary statistics.
5. PRSFNN Snakemake [workflow](https://github.com/weinstockj/PRSFNN-runner).
6. [Script](https://github.com/weinstockj/download_high_depth_1kg) to download 1000 Genomes 30x data.
7. [Scripts](https://github.com/weinstockj/dbSNP_to_parquet) to create rsid lookup tables from dbSNP.

## 6 Data Availability

## 7 Author Contributions

Joshua Weinstock: Conceptualization, Data curation, Methodology, Software, Validation, Writing - original draft, Writing - review & editing April Kim: Data curation, Formal analysis, Methodology, Software, Validation, Writing - original draft, Writing - review & editing Alexis Battle: Conceptualization, Resources, Supervision, Writing - review & editing

## 8 Acknowledgments

We thank Marios Arvanitis, Ashton Omdahl, Seraj Grimes, Sohail Zahid, and David Cutler for helpful discussions. The authors would like to thank Battle lab members for helpful discussion throughout the course of this work.

## Appendix A Effect of LD reference panel on SBayesRC performance

**Fig. A1.**
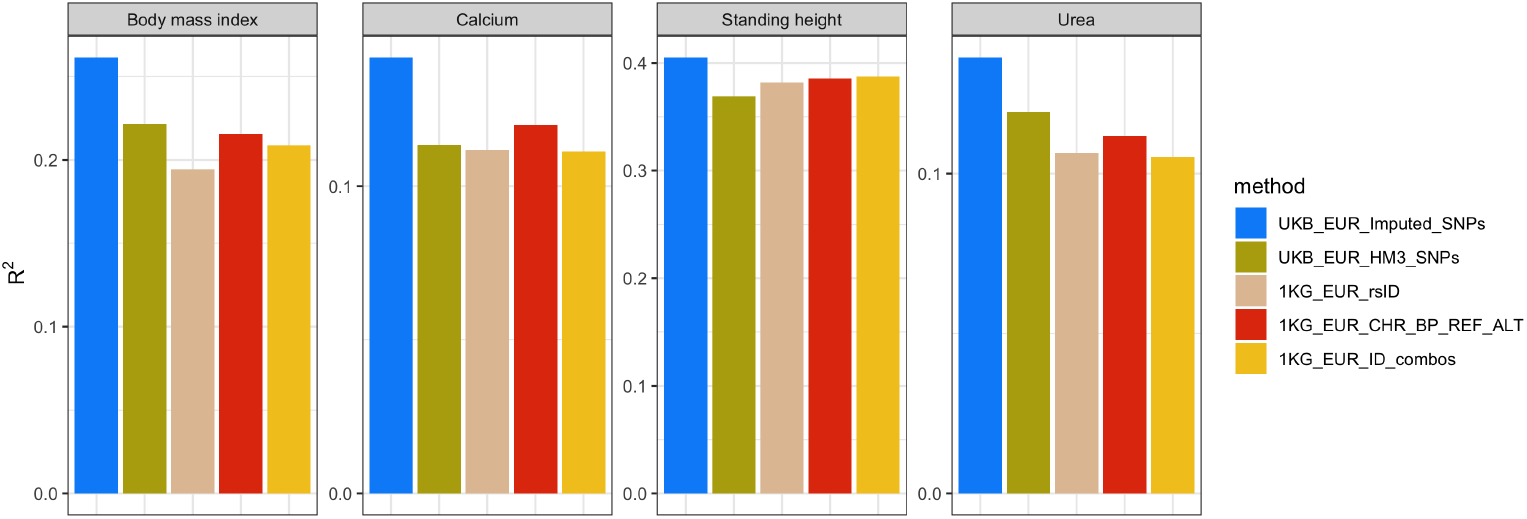
Sensitivity of prediction performance to the choice of LD reference.

We observed that SBayesRC exhibited reduced predictive accuracy when using the 1kGP as the LD reference panel. To investigate this observation, we considered prior literature assessing the sensitivity of polygenic risk scores (PRS) methods to the choice of LD reference.

The authors of SBayesRC has evaluated its robustness to different LD reference panels under various simulation scenarios [61]. In particular, they compared prediction accuracy when using LD panels derived from the 1kGP (specifically, a European subset of 500 individuals referred to as “1kg0.5k”), UKB (20,000 random individuals), and UK10K (3,642 unrelated individuals). Their results show that methods including LDpred2-inf, SBayesR, and SBayesRC all experienced diminished performance when relying on the smaller 1kGP panel. These findings support the known limitation that PRS methods often suffer from reduced imputation accuracy and convergence issues when the LD reference lacks sufficient sample size or population match.

In a separate, recently published study [46], the authors performed a sensitivity analysis on the choice of LD reference panel using height and BMI GWAS summary statistics from the GIANT consortium. They demonstrated that switching from the 1kGP LD panel to the UKB LD panel substantially improved PRS accuracy across all evaluated methods. Notably, SBayesRC was only evaluated using UKB LD panels in that study, since SBayesRC does not provide LD data for the 1kGP reference. This omission left a gap in comparative benchmarking, which our analysis addresses.

To ensure that the observed performance gap was not attributable to implementation/technical errors, we evaluated alternative SNP identifier harmonization strategies. The baseline annotation and LD panel files for SBayesRC uses rsIDs, and our initial approach matched the LD reference and annotation SNP identifiers accordingly. Given the risk of mapping errors (e.g., multiple rsIDs per position, outdated identifiers), we adopted an alternative scheme that reformatted both the LD reference and annotation files using `CHR_BP_REF_ALT` identifiers. Although this reduced the number of SNPs in the annotation set (∼600K SNPs excluded from ∼8.2M total), it ensured identifier consistency and maximal overlap across inputs.

We further created a hybrid identifier scheme wherein SNPs with resolvable rsIDs were retained, while unmatched SNPs preserved their `CHR_BP_REF_ALT` identifiers. Across these harmonization strategies, we observed modest improvements in PRS performance (Supplementary Fig. 1), suggesting that inconsistent identifiers partially contributed to the reduced accuracy.

Together, these findings indicate that the reduced performance of SBayesRC with 1kGP-based LD reference panels is primarily due to the limited sample size and lower SNP overlap with GWAS and annotation inputs.

## Appendix B CPU hours and user run time of PRS

**Fig. B2.**
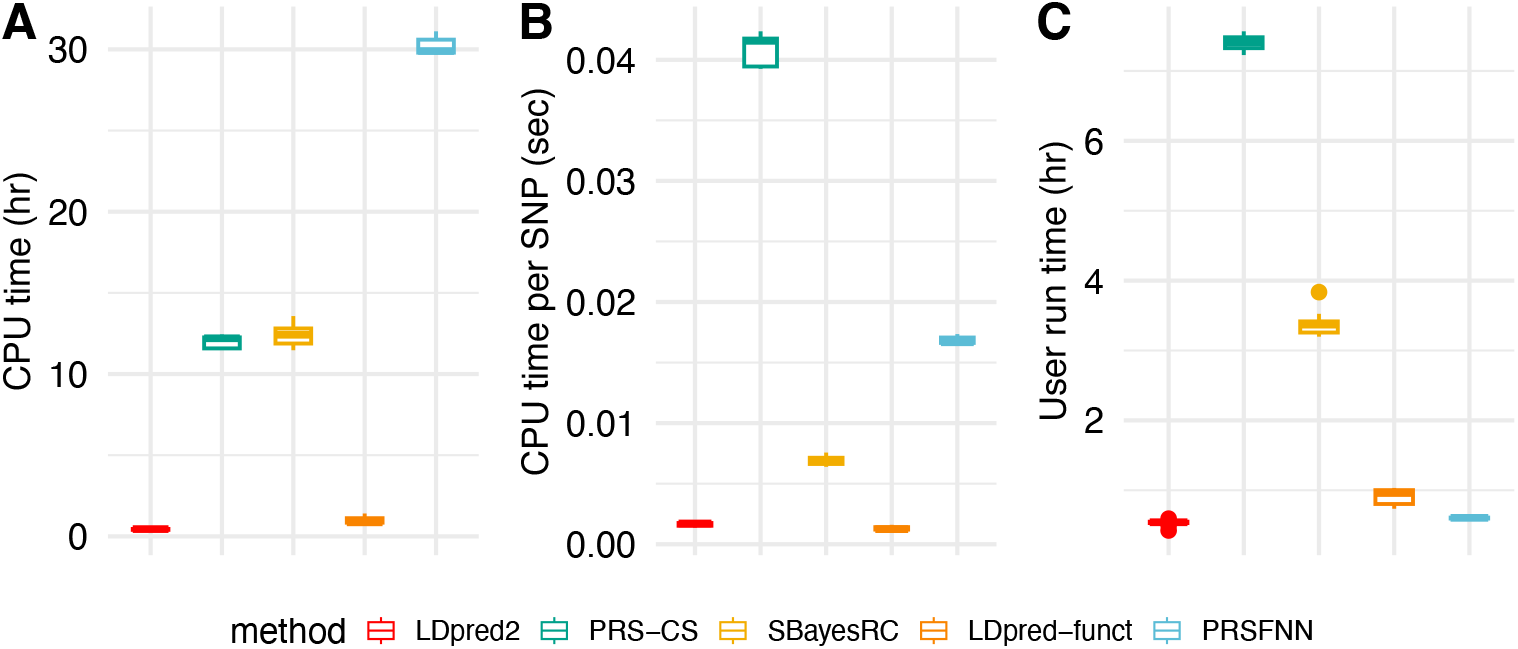
The total user and system CPU hours and user run time of 5 PRS methods reported. Each boxplot shows the distribution of the CPU hours and user run time for a given method based on 10 independent runs across 10 phenotypes.

## Appendix C Post-training model interpretation – enrichment analysis

**Fig. C3.**
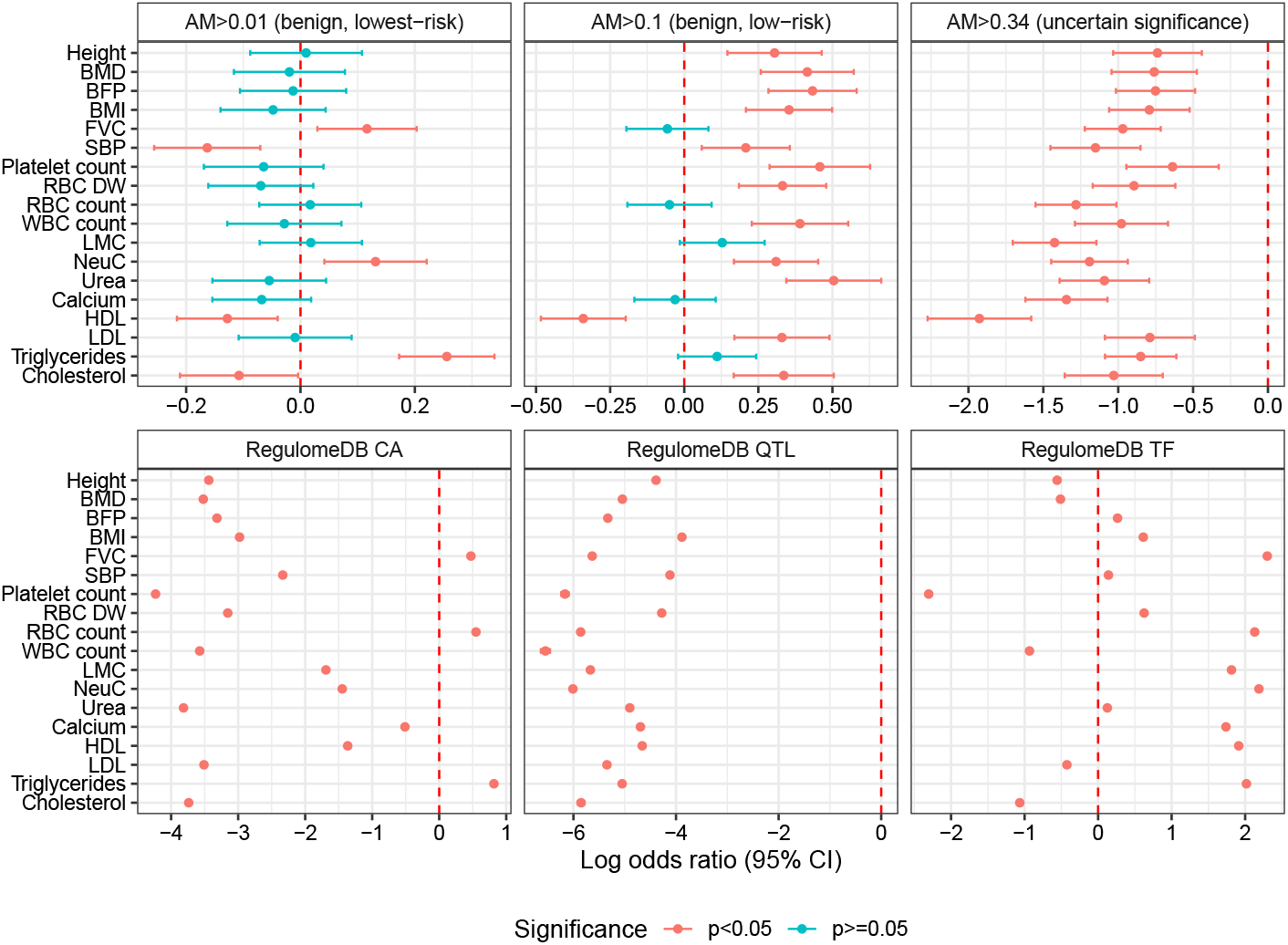
Logistic regression model coefficients – RegulomeDB and AlphaMissense

**Table C1.** Logistic regression model for height.

|  | *Dependent variable:* |
| --- | --- |
|  | Favored by NN |
| number of accessible cell types $\geq 5$ | 1.22***<br>(0.01) |
| number of accessible cell types $\geq 10$ | 1.31***<br>(0.02) |
| number of accessible cell types $\geq 20$ | 1.78***<br>(0.02) |
| number of accessible cell types $\geq 50$ | -1.27***<br>(0.02) |
| TF PWM | -4.96***<br>(0.02) |
| RegulomeDB TF | -0.56***<br>(0.01) |
| RegulomeDB CA | -3.44***<br>(0.01) |
| ENCODE4 CA | -7.71***<br>(0.27) |
| Zoonomia RoCC | -2.37***<br>(0.05) |
| ENCODE4 promoter-like | -1.49***<br>(0.03) |
| ENCODE4 proximal enhancer-like | -0.51***<br>(0.02) |
| RegulomeDB RegulomeDB QTL | -4.39***<br>(0.02) |
| ENCODE4 distal enhancer-like | -0.48***<br>(0.01) |
| ENCODE4 CA+TF | -3.96***<br>(0.14) |
| ENCODE4 TF | -6.71***<br>(0.24) |
| ENCODE4 CA+CTCF | -5.19***<br>(0.11) |
| ENCODE4 CA+H3K4me3 | -4.77***<br>(0.10) |
| 1kGP MAF Rare | 3.58***<br>(0.004) |
| AlphaMissense spline 1 | 0.01<br>(0.05) |
| AlphaMissense spline 2 | 0.30***<br>(0.08) |
| AlphaMissense spline 3 | -0.74***<br>(0.15) |
| Constant | -2.66***<br>(0.003) |
| Observations | 6,453,934 |
| Log Likelihood | -983,362.70 |
| Akaike Inf. Crit. | 1,966,769.00 |
| Note: | *p<0.1; **p<0.05; ***p<0.01 |

**Table C2.** Logistic regression model for bone mineral density.

|  | *Dependent variable:* |
| --- | --- |
|  | Favored by NN |
| number of accessible cell types $\geq 5$ | 1.84***<br>(0.01) |
| number of accessible cell types $\geq 10$ | 1.11***<br>(0.02) |
| number of accessible cell types $\geq 20$ | 1.01***<br>(0.02) |
| number of accessible cell types $\geq 50$ | -0.69***<br>(0.02) |
| TF PWM | -5.48***<br>(0.03) |
| RegulomeDB TF | -0.51***<br>(0.01) |
| RegulomeDB CA | -3.52***<br>(0.01) |
| ENCODE4 CA | -7.03***<br>(0.19) |
| Zoonomia RoCC | -1.82***<br>(0.04) |
| ENCODE4 promoter-like | -0.50***<br>(0.03) |
| ENCODE4 proximal enhancer-like | -0.39***<br>(0.02) |
| RegulomeDB RegulomeDB QTL | -5.05***<br>(0.02) |
| ENCODE4 distal enhancer-like | -0.55***<br>(0.01) |
| ENCODE4 CA+TF | -3.42***<br>(0.11) |
| ENCODE4 TF | -5.92***<br>(0.16) |
| ENCODE4 CA+CTCF | -4.55***<br>(0.08) |
| ENCODE4 CA+H3K4me3 | -4.27***<br>(0.08) |
| 1kGP MAF Rare | 3.66***<br>(0.004) |
| AlphaMissense spline 1 | -0.02<br>(0.05) |
| AlphaMissense spline 2 | 0.42***<br>(0.08) |
| AlphaMissense spline 3 | -0.76***<br>(0.15) |
| Constant | -2.73***<br>(0.003) |
| Observations | 6,453,269 |
| Log Likelihood | -957,350.70 |
| Akaike Inf. Crit. | 1,914,745.00 |
| *Note:* | *p<0.1; **p<0.05; ***p<0.01 |

**Table C3.** Logistic regression model for body fat percentage.

|  | *Dependent variable:* |
| --- | --- |
|  | Favored by NN |
| number of accessible cell types $\geq 5$ | 1.88***<br>(0.01) |
| number of accessible cell types $\geq 10$ | 0.66***<br>(0.01) |
| number of accessible cell types $\geq 20$ | 0.73***<br>(0.02) |
| number of accessible cell types $\geq 50$ | 0.30***<br>(0.02) |
| TF PWM | -5.69***<br>(0.03) |
| RegulomeDB TF | 0.26***<br>(0.01) |
| RegulomeDB CA | -3.32***<br>(0.01) |
| ENCODE4 CA | -6.33***<br>(0.15) |
| Zoonomia RoCC | -1.48***<br>(0.04) |
| ENCODE4 promoter-like | -0.03<br>(0.03) |
| ENCODE4 proximal enhancer-like | 0.23***<br>(0.01) |
| RegulomeDB RegulomeDB QTL | -5.33***<br>(0.02) |
| ENCODE4 distal enhancer-like | 0.19***<br>(0.01) |
| ENCODE4 CA+TF | -3.06***<br>(0.09) |
| ENCODE4 TF | -5.71***<br>(0.14) |
| ENCODE4 CA+CTCF | -3.92***<br>(0.06) |
| ENCODE4 CA+H3K4me3 | -3.88***<br>(0.07) |
| 1kGP MAF Rare | 3.69***<br>(0.004) |
| AlphaMissense spline 1 | -0.01<br>(0.05) |
| AlphaMissense spline 2 | 0.43***<br>(0.08) |
| AlphaMissense spline 3 | -0.75***<br>(0.13) |
| Constant | -2.98***<br>(0.003) |
| Observations | 6,453,865 |
| Log Likelihood | -969,820.80 |
| Akaike Inf. Crit. | 1,939,686.00 |
| *Note:* | *p<0.1; **p<0.05; ***p<0.01 |

**Table C4.**
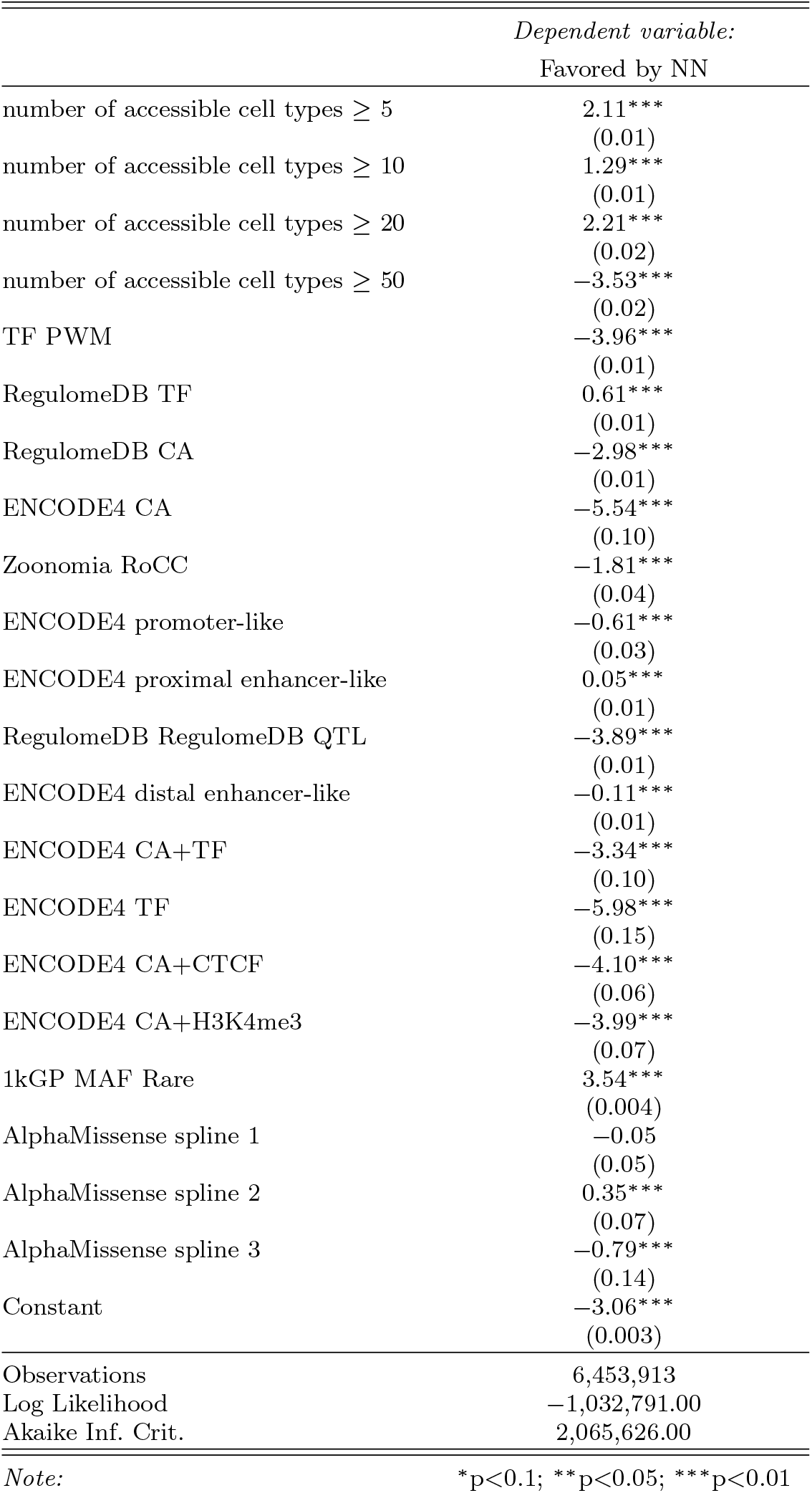
Logisitc regression model for body mass index.

**Table C5.**
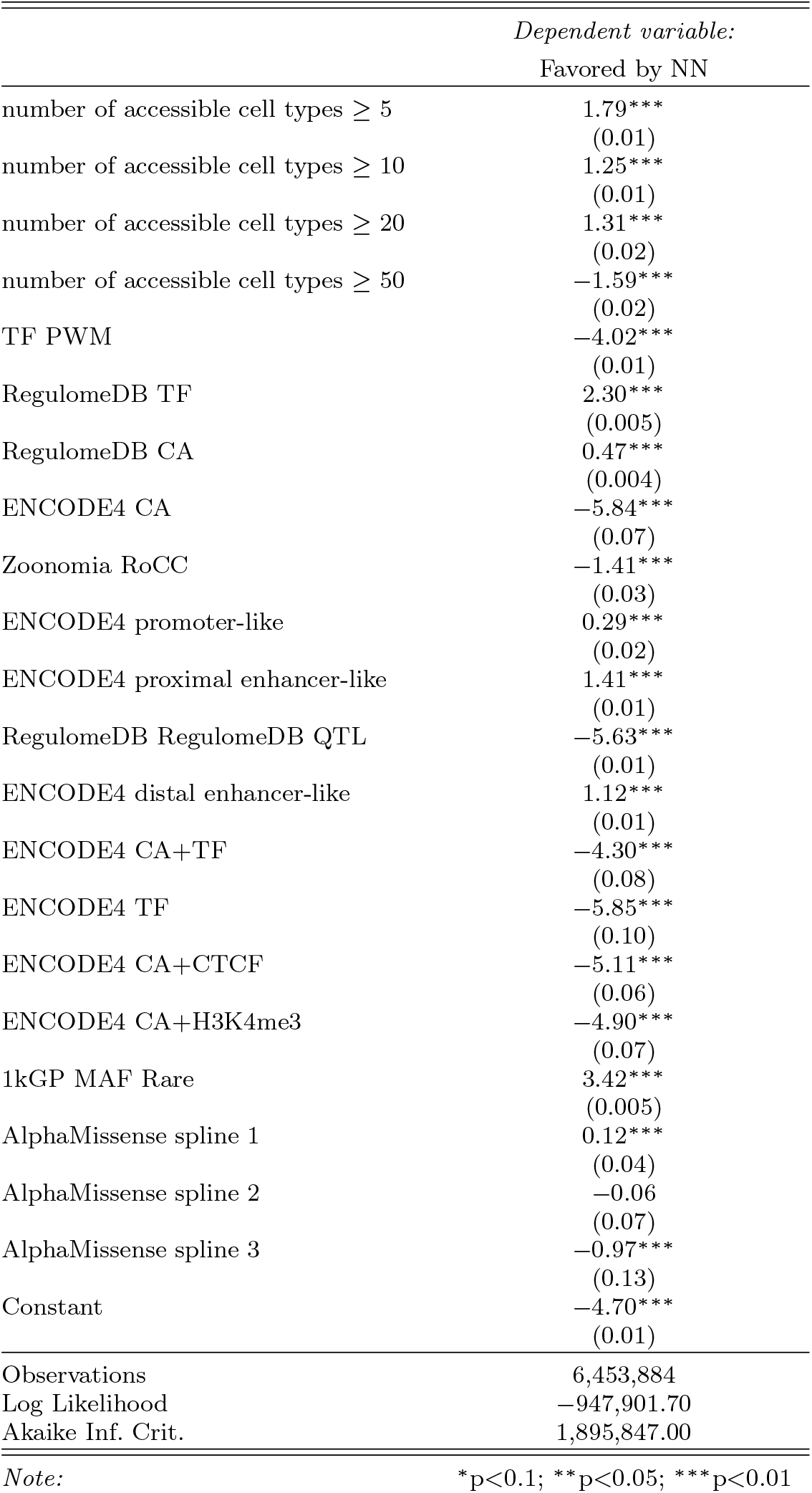
Logistic regression model for forced vital capacity.

**Table C6.** Logistic regression model for systolic blood pressure.

|  | *Dependent variable:* |  |
| --- | --- | --- |
|  | Favored by NN |  |
| number of accessible cell types $\geq 5$ | 1.37***<br>(0.01) | |
| number of accessible cell types $\geq 10$ | 1.00***<br>(0.01) | |
| number of accessible cell types $\geq 20$ | 1.72***<br>(0.02) | |
| number of accessible cell types $\geq 50$ | -1.63***<br>(0.02) | |
| TF PWM | -4.18***<br>(0.01) |  |
| RegulomeDB TF | 0.14***<br>(0.01) |  |
| RegulomeDB CA | -2.33***<br>(0.01) |  |
| ENCODE4 CA | -7.66***<br>(0.28) |  |
| Zoonomia RoCC | -2.27***<br>(0.05) |  |
| ENCODE4 promoter-like | -1.34***<br>(0.03) |  |
| ENCODE4 proximal enhancer-like | -0.46***<br>(0.01) |  |
| RegulomeDB RegulomeDB QTL | -4.12***<br>(0.01) |  |
| ENCODE4 distal enhancer-like | -0.11***<br>(0.01) |  |
| ENCODE4 CA+TF | -4.02***<br>(0.14) |  |
| ENCODE4 TF | -7.63***<br>(0.38) |  |
| ENCODE4 CA+CTCF | -4.91***<br>(0.10) |  |
| ENCODE4 CA+H3K4me3 | -4.88***<br>(0.11) |  |
| 1kGP MAF Rare | 3.61***<br>(0.004) |  |
| AlphaMissense spline 1 | -0.16***<br>(0.05) |  |
| AlphaMissense spline 2 | 0.21***<br>(0.08) |  |
| AlphaMissense spline 3 | -1.15***<br>(0.15) |  |
| Constant | -3.01***<br>(0.003) |  |
| Observations | 6,454,571 |  |
| Log Likelihood | -1,054,017.00 |  |
| Akaike Inf. Crit. | 2,108,078.00 |  |
| *Note:* | *p<0.1; **p<0.05; ***p<0.01 |  |

**Table C7.** Logistic regression model for platelet count.

|  | *Dependent variable:* |
| --- | --- |
|  | Favored by NN |
| number of accessible cell types $\geq 5$ | 0.64***<br>(0.02) |
| number of accessible cell types $\geq 10$ | 0.75***<br>(0.02) |
| number of accessible cell types $\geq 20$ | 1.21***<br>(0.02) |
| number of accessible cell types $\geq 50$ | 1.73***<br>(0.03) |
| TF PWM | -7.36***<br>(0.06) |
| RegulomeDB TF | -2.30***<br>(0.01) |
| RegulomeDB CA | -4.23***<br>(0.01) |
| ENCODE4 CA | -8.87***<br>(0.44) |
| Zoonomia RoCC | -1.97***<br>(0.05) |
| ENCODE4 promoter-like | -1.10***<br>(0.03) |
| ENCODE4 proximal enhancer-like | -0.67***<br>(0.02) |
| RegulomeDB QTL | -6.17***<br>(0.04) |
| ENCODE4 distal enhancer-like | -0.85***<br>(0.01) |
| ENCODE4 CA+TF | -3.66***<br>(0.12) |
| ENCODE4 TF | -6.89***<br>(0.27) |
| ENCODE4 CA+CTCF | -5.06***<br>(0.10) |
| ENCODE4 CA+H3K4me3 | -4.79***<br>(0.10) |
| 1kGP MAF Rare | 3.58***<br>(0.004) |
| AlphaMissense spline 1 | -0.06<br>(0.05) |
| AlphaMissense spline 2 | 0.46***<br>(0.09) |
| AlphaMissense spline 3 | -0.64***<br>(0.16) |
| Constant | -2.32***<br>(0.003) |
| Observations | 6,453,938 |
| Log Likelihood | -920,479.60 |
| Akaike Inf. Crit. | 1,841,003.00 |
| *Note:* | *p<0.1; **p<0.05; ***p<0.01 |

**Table C8.** Logistic regression model for red blood cell distribution width.

|  | *Dependent variable:* |
| --- | --- |
|  | Favored by NN |
| number of accessible cell types $\geq 5$ | 1.49***<br>(0.01) |
| number of accessible cell types $\geq 10$ | 0.82***<br>(0.01) |
| number of accessible cell types $\geq 20$ | 1.55***<br>(0.02) |
| number of accessible cell types $\geq 50$ | -1.11***<br>(0.02) |
| TF PWM | -4.48***<br>(0.02) |
| RegulomeDB TF | 0.63***<br>(0.01) |
| RegulomeDB CA | -3.16***<br>(0.01) |
| ENCODE4 CA | -7.24***<br>(0.24) |
| Zoonomia RoCC | -2.01***<br>(0.04) |
| ENCODE4 promoter-like | -0.79***<br>(0.03) |
| ENCODE4 proximal enhancer-like | 0.37***<br>(0.01) |
| RegulomeDB QTL | -4.28***<br>(0.01) |
| ENCODE4 distal enhancer-like | 0.24***<br>(0.01) |
| ENCODE4 CA+TF | -3.74***<br>(0.12) |
| ENCODE4 TF | -6.80***<br>(0.22) |
| ENCODE4 CA+CTCF | -4.49***<br>(0.08) |
| ENCODE4 CA+H3K4me3 | -4.29***<br>(0.09) |
| 1kGP MAF Rare | 3.61***<br>(0.004) |
| AlphaMissense spline 1 | -0.07<br>(0.05) |
| AlphaMissense spline 2 | 0.33***<br>(0.08) |
| AlphaMissense spline 3 | -0.89***<br>(0.14) |
| Constant | -3.03***<br>(0.003) |
| Observations | 6,453,939 |
| Log Likelihood | -1,017,515.00 |
| Akaike Inf. Crit. | 2,035,074.00 |
| *Note:* *p<0.1; **p<0.05; ***p<0.01 |  |

**Table C9.**
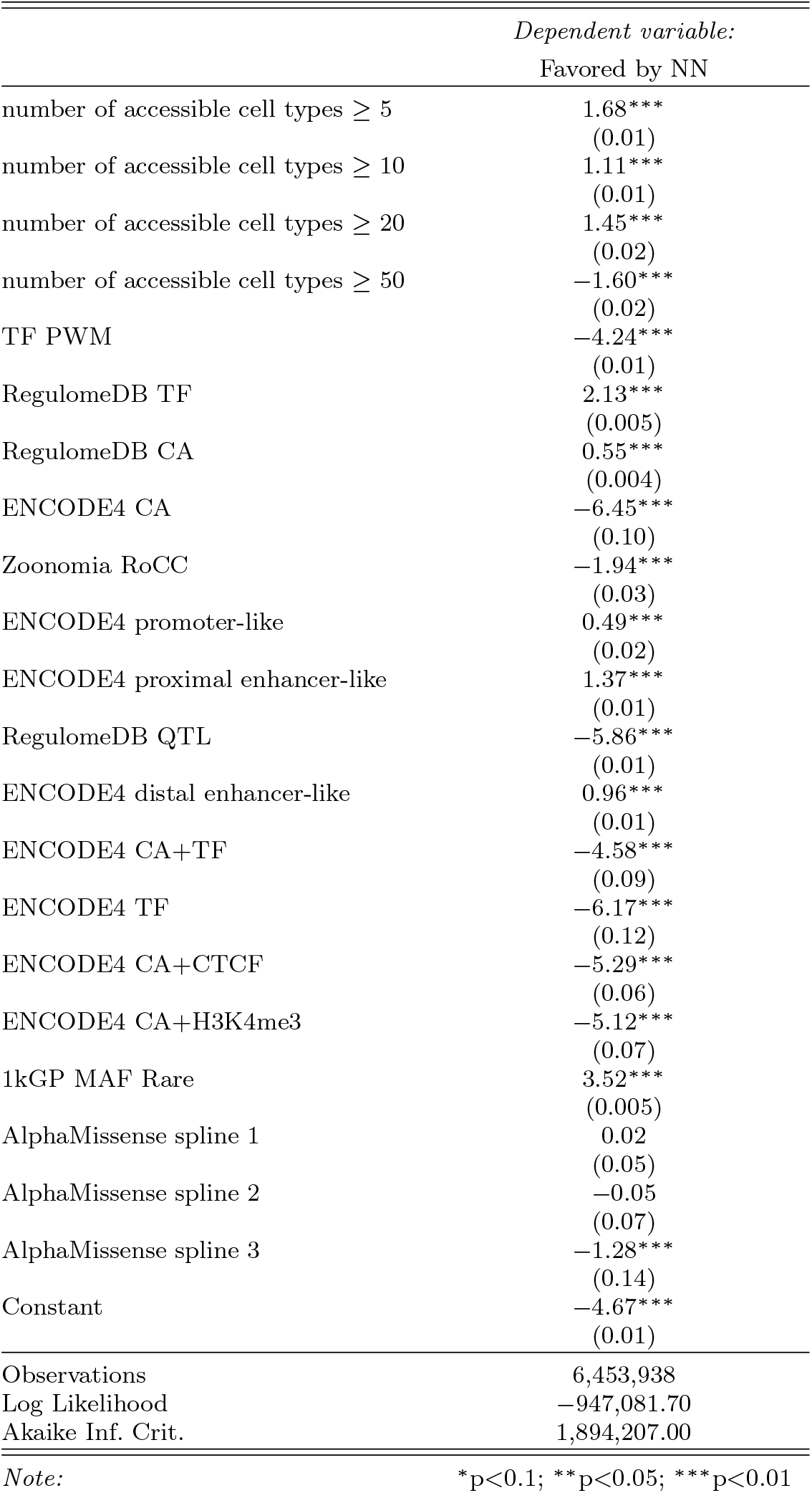
Logistic regression model for red blood cell count.

**Table C10.** Logistic regression model for white blood cell count.

|  | *Dependent variable:* |
| --- | --- |
|  | Favored by NN |
| number of accessible cell types $\geq 5$ | 1.69***<br>(0.01) |
| number of accessible cell types $\geq 10$ | 1.30***<br>(0.02) |
| number of accessible cell types $\geq 20$ | 0.73***<br>(0.02) |
| number of accessible cell types $\geq 50$ | -0.82***<br>(0.02) |
| TF PWM | -6.43***<br>(0.04) |
| RegulomeDB TF | -0.93***<br>(0.01) |
| RegulomeDB CA | -3.57***<br>(0.01) |
| ENCODE4 CA | -7.44***<br>(0.22) |
| Zoonomia RoCC | -2.51***<br>(0.05) |
| ENCODE4 promoter-like | -0.74***<br>(0.03) |
| ENCODE4 proximal enhancer-like | -0.52***<br>(0.02) |
| RegulomeDB QTL | -6.55***<br>(0.05) |
| ENCODE4 distal enhancer-like | -0.75***<br>(0.01) |
| ENCODE4 CA+TF | -3.81***<br>(0.12) |
| ENCODE4 TF | -6.51***<br>(0.22) |
| ENCODE4 CA+CTCF | -5.25***<br>(0.11) |
| ENCODE4 CA+H3K4me3 | -4.61***<br>(0.09) |
| 1kGP MAF Rare | 3.65***<br>(0.004) |
| AlphaMissense spline 1 | -0.03<br>(0.05) |
| AlphaMissense spline 2 | 0.39***<br>(0.08) |
| AlphaMissense spline 3 | -0.98***<br>(0.16) |
| Constant | -2.60***<br>(0.003) |
| Observations | 6,453,941 |
| Log Likelihood | -944,915.50 |
| Akaike Inf. Crit. | 1,889,875.00 |
| *Note:* *p<0.1; **p<0.05; ***p<0.01 |  |

**Table C11.** Logistic regression model for lymphocyte count.

|  | *Dependent variable:* |
| --- | --- |
|  | Favored by NN |
| number of accessible cell types $\geq 5$ | 1.20***<br>(0.01) |
| number of accessible cell types $\geq 10$ | 0.69***<br>(0.02) |
| number of accessible cell types $\geq 20$ | 1.00***<br>(0.02) |
| number of accessible cell types $\geq 50$ | 1.17***<br>(0.02) |
| TF PWM | -5.29***<br>(0.02) |
| RegulomeDB TF | 1.81***<br>(0.01) |
| RegulomeDB CA | -1.69***<br>(0.01) |
| ENCODE4 CA | -8.99***<br>(0.58) |
| Zoonomia RoCC | -2.51***<br>(0.04) |
| ENCODE4 promoter-like | 0.02<br>(0.03) |
| ENCODE4 proximal enhancer-like | 1.57***<br>(0.01) |
| RegulomeDB QTL | -5.67***<br>(0.02) |
| ENCODE4 distal enhancer-like | 1.42***<br>(0.01) |
| ENCODE4 CA+TF | -4.66***<br>(0.16) |
| ENCODE4 TF | -7.25***<br>(0.24) |
| ENCODE4 CA+CTCF | -4.58***<br>(0.08) |
| ENCODE4 CA+H3K4me3 | -4.27***<br>(0.08) |
| 1kGP MAF Rare | 4.18***<br>(0.01) |
| AlphaMissense spline 1 | 0.02<br>(0.05) |
| AlphaMissense spline 2 | 0.13*<br>(0.07) |
| AlphaMissense spline 3 | -1.43***<br>(0.14) |
| Constant | -4.29***<br>(0.01) |
| Observations | 6,453,938 |
| Log Likelihood | -928,238.50 |
| Akaike Inf. Crit. | 1,856,521.00 |
| *Note:* | *p<0.1; **p<0.05; ***p<0.01 |

**Table C12.**
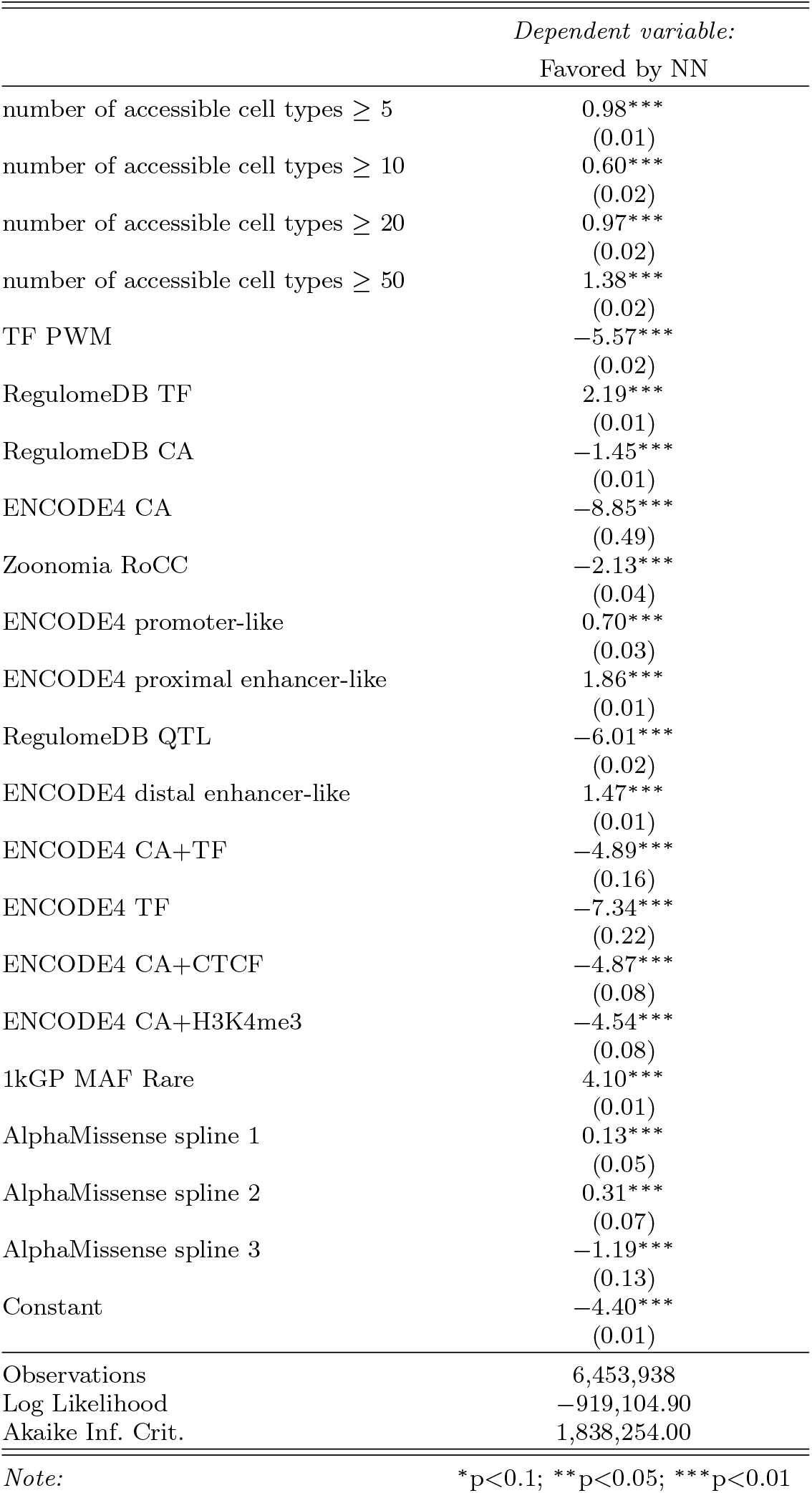
Logistic regression model for neutrophil count.

**Table C13.**
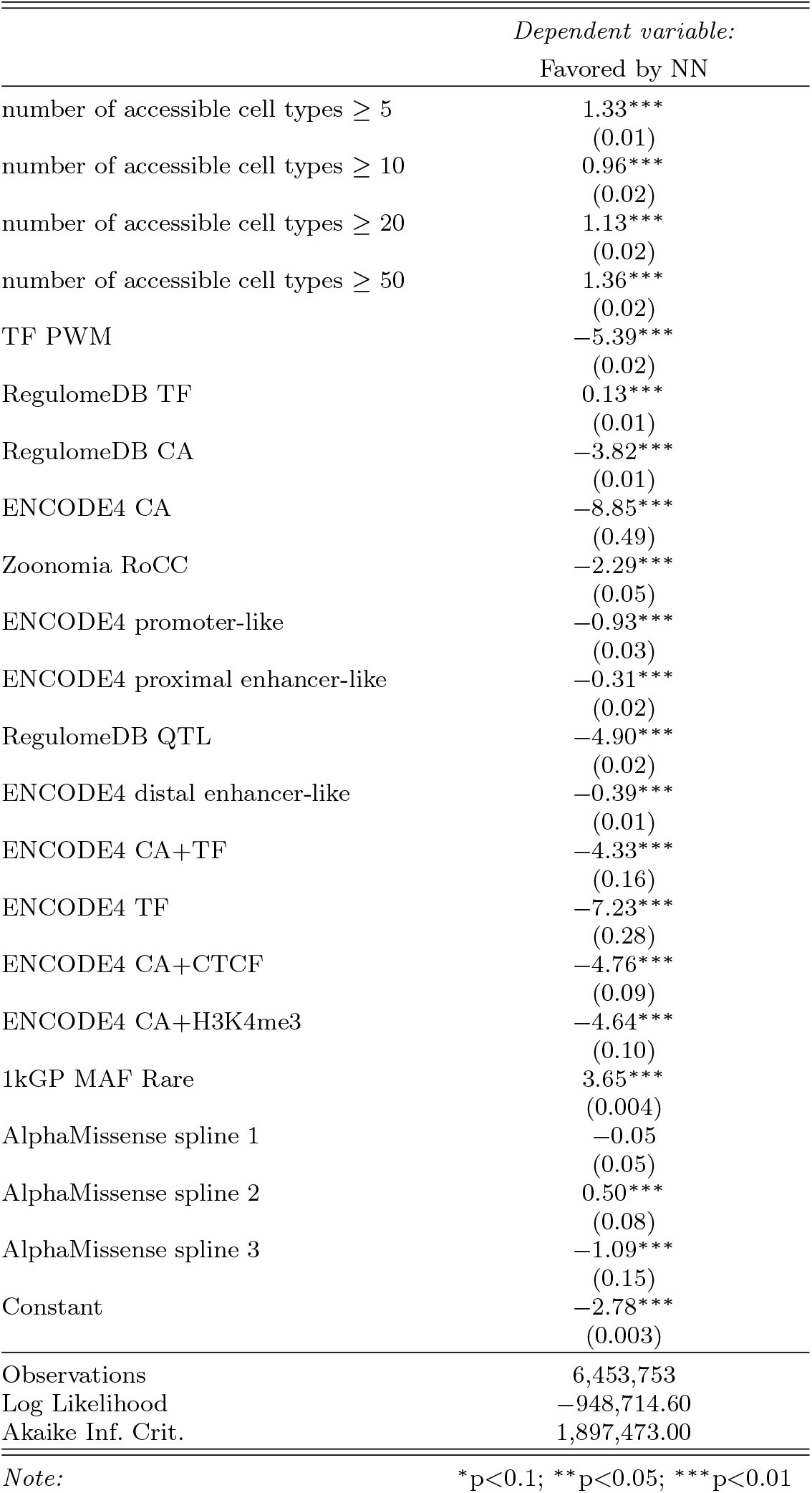
Logistic regression model for urea.

|  | *Dependent variable:* |
| --- | --- |
|  | Favored by NN |
| number of accessible cell types $\geq 5$ | 1.33***<br>(0.01) |
| number of accessible cell types $\geq 10$ | 0.96***<br>(0.02) |
| number of accessible cell types $\geq 20$ | 1.13***<br>(0.02) |
| number of accessible cell types $\geq 50$ | 1.36***<br>(0.02) |
| TF PWM | -5.39***<br>(0.02) |
| RegulomeDB TF | 0.13***<br>(0.01) |
| RegulomeDB CA | -3.82***<br>(0.01) |
| ENCODE4 CA | -8.85***<br>(0.49) |
| Zoonomia RoCC | -2.29***<br>(0.05) |
| ENCODE4 promoter-like | -0.93***<br>(0.03) |
| ENCODE4 proximal enhancer-like | -0.31***<br>(0.02) |
| RegulomeDB QTL | -4.90***<br>(0.02) |
| ENCODE4 distal enhancer-like | -0.39***<br>(0.01) |
| ENCODE4 CA+TF | -4.33***<br>(0.16) |
| ENCODE4 TF | -7.23***<br>(0.28) |
| ENCODE4 CA+CTCF | -4.76***<br>(0.09) |
| ENCODE4 CA+H3K4me3 | -4.64***<br>(0.10) |
| 1kGP MAF Rare | 3.65***<br>(0.004) |
| AlphaMissense spline 1 | -0.05<br>(0.05) |
| AlphaMissense spline 2 | 0.50***<br>(0.08) |
| AlphaMissense spline 3 | -1.09***<br>(0.15) |
| Constant | -2.78***<br>(0.003) |
| Observations | 6,453,753 |
| Log Likelihood | -948,714.60 |
| Akaike Inf. Crit. | 1,897,473.00 |
| *Note:* | *p<0.1; **p<0.05; ***p<0.01 |

**Table C14.**
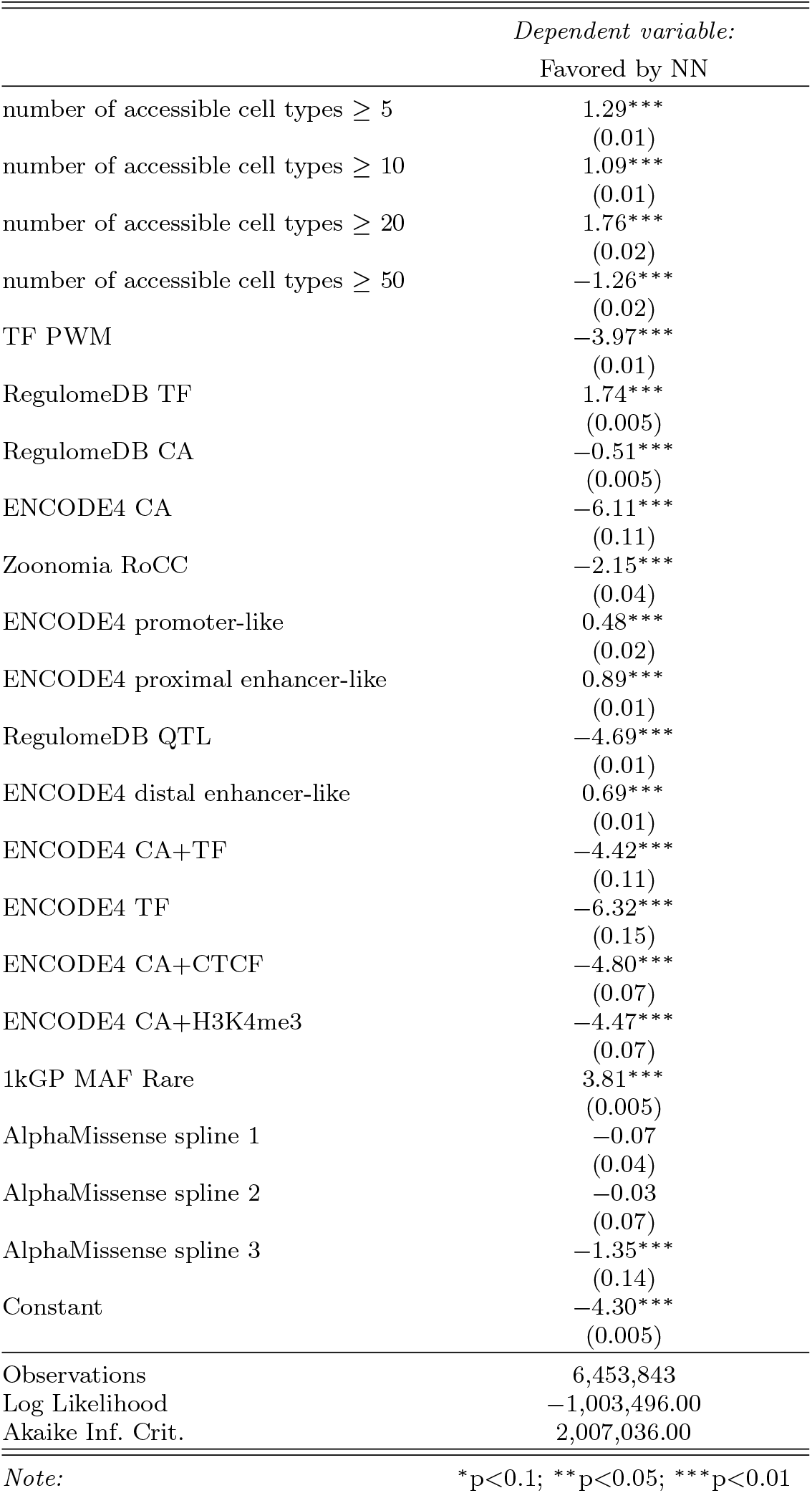
Logistic regression model for calcium.

**Table C15.** Logistic regression model for HDL.

|  | *Dependent variable:* |
| --- | --- |
|  | Favored by NN |
| number of accessible cell types $\geq 5$ | 2.01***<br>(0.01) |
| number of accessible cell types $\geq 10$ | 1.25***<br>(0.01) |
| number of accessible cell types $\geq 20$ | 1.69***<br>(0.02) |
| number of accessible cell types $\geq 50$ | -2.31***<br>(0.02) |
| TF PWM | -3.85***<br>(0.01) |
| RegulomeDB TF | 1.91***<br>(0.01) |
| RegulomeDB CA | -1.37***<br>(0.01) |
| ENCODE4 CA | -6.02***<br>(0.11) |
| Zoonomia RoCC | -2.11***<br>(0.04) |
| ENCODE4 promoter-like | -0.68***<br>(0.03) |
| ENCODE4 proximal enhancer-like | 0.50***<br>(0.01) |
| RegulomeDB QTL | -4.66***<br>(0.01) |
| ENCODE4 distal enhancer-like | 0.48***<br>(0.01) |
| ENCODE4 CA+TF | -4.17***<br>(0.10) |
| ENCODE4 TF | -6.83***<br>(0.18) |
| ENCODE4 CA+CTCF | -4.71***<br>(0.07) |
| ENCODE4 CA+H3K4me3 | -4.66***<br>(0.08) |
| 1kGP MAF Rare | 4.04***<br>(0.01) |
| AlphaMissense spline 1 | -0.13***<br>(0.04) |
| AlphaMissense spline 2 | -0.34***<br>(0.07) |
| AlphaMissense spline 3 | -1.93***<br>(0.18) |
| Constant | -4.27***<br>(0.005) |
| Observations | 6,453,825 |
| Log Likelihood | -978,482.10 |
| Akaike Inf. Crit. | 1,957,008.00 |
| *Note:* | *p<0.1; **p<0.05; ***p<0.01 |

**Table C16.** Logistic regression model for LDL.

|  | *Dependent variable:* |
| --- | --- |
|  | Favored by NN |
| number of accessible cell types $\geq 5$ | 1.83***<br>(0.01) |
| number of accessible cell types $\geq 10$ | 1.44***<br>(0.02) |
| number of accessible cell types $\geq 20$ | 1.06***<br>(0.02) |
| number of accessible cell types $\geq 50$ | -2.34***<br>(0.02) |
| TF PWM | -5.20***<br>(0.02) |
| RegulomeDB TF | -0.42***<br>(0.01) |
| RegulomeDB CA | -3.51***<br>(0.01) |
| ENCODE4 CA | -7.04***<br>(0.19) |
| Zoonomia RoCC | -2.34***<br>(0.05) |
| ENCODE4 promoter-like | -0.67***<br>(0.03) |
| ENCODE4 proximal enhancer-like | -0.21***<br>(0.02) |
| RegulomeDB QTL | -5.35***<br>(0.03) |
| ENCODE4 distal enhancer-like | -0.59***<br>(0.01) |
| ENCODE4 CA+TF | -3.50***<br>(0.11) |
| ENCODE4 TF | -6.08***<br>(0.17) |
| ENCODE4 CA+CTCF | -4.66***<br>(0.08) |
| ENCODE4 CA+H3K4me3 | -4.59***<br>(0.09) |
| 1kGP MAF Rare | 3.65***<br>(0.004) |
| AlphaMissense spline 1 | -0.01<br>(0.05) |
| AlphaMissense spline 2 | 0.33***<br>(0.08) |
| AlphaMissense spline 3 | -0.79***<br>(0.15) |
| Constant | -2.73***<br>(0.003) |
| Observations | 6,453,770 |
| Log Likelihood | -957,161.30 |
| Akaike Inf. Crit. | 1,914,367.00 |
| *Note:* *p<0.1; **p<0.05; ***p<0.01 |  |

**Table C17.** Logistic regression model for triglycerides.

|  | *Dependent variable:* |
| --- | --- |
|  | Favored by NN |
| number of accessible cell types $\geq 5$ | 1.50***<br>(0.01) |
| number of accessible cell types $\geq 10$ | 1.30***<br>(0.01) |
| number of accessible cell types $\geq 20$ | 1.88***<br>(0.02) |
| number of accessible cell types $\geq 50$ | -2.45***<br>(0.02) |
| TF PWM | -3.86***<br>(0.01) |
| RegulomeDB TF | 2.02***<br>(0.005) |
| RegulomeDB CA | 0.82***<br>(0.004) |
| ENCODE4 CA | -5.81***<br>(0.07) |
| Zoonomia RoCC | -1.45***<br>(0.03) |
| ENCODE4 promoter-like | 0.57***<br>(0.02) |
| ENCODE4 proximal enhancer-like | 1.32***<br>(0.01) |
| RegulomeDB QTL | -5.05***<br>(0.01) |
| ENCODE4 distal enhancer-like | 0.66***<br>(0.01) |
| ENCODE4 CA+TF | -4.25***<br>(0.08) |
| ENCODE4 TF | -5.69***<br>(0.10) |
| ENCODE4 CA+CTCF | -4.81***<br>(0.05) |
| ENCODE4 CA+H3K4me3 | -4.68***<br>(0.06) |
| 1kGP MAF Rare | 3.35***<br>(0.005) |
| AlphaMissense spline 1 | 0.26***<br>(0.04) |
| AlphaMissense spline 2 | 0.11*<br>(0.07) |
| AlphaMissense spline 3 | -0.85***<br>(0.12) |
| Constant | -4.61***<br>(0.005) |
| Observations | 6,453,761 |
| Log Likelihood | -981,913.00 |
| Akaike Inf. Crit. | 1,963,870.00 |
| Note: | *p<0.1; **p<0.05; ***p<0.01 |

**Table C18.**
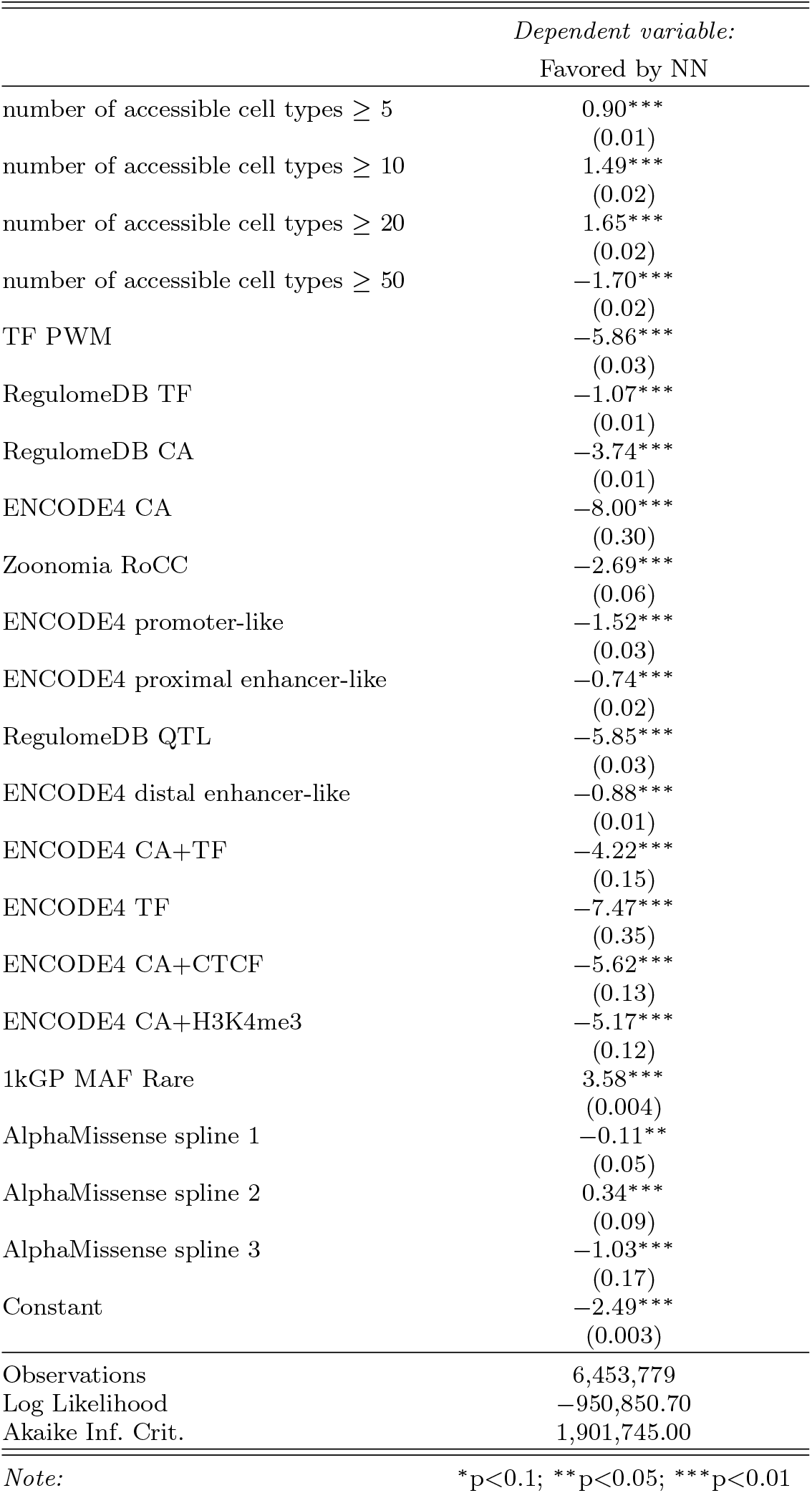
Logistic regression model for cholesterol.

**Fig. C4.**
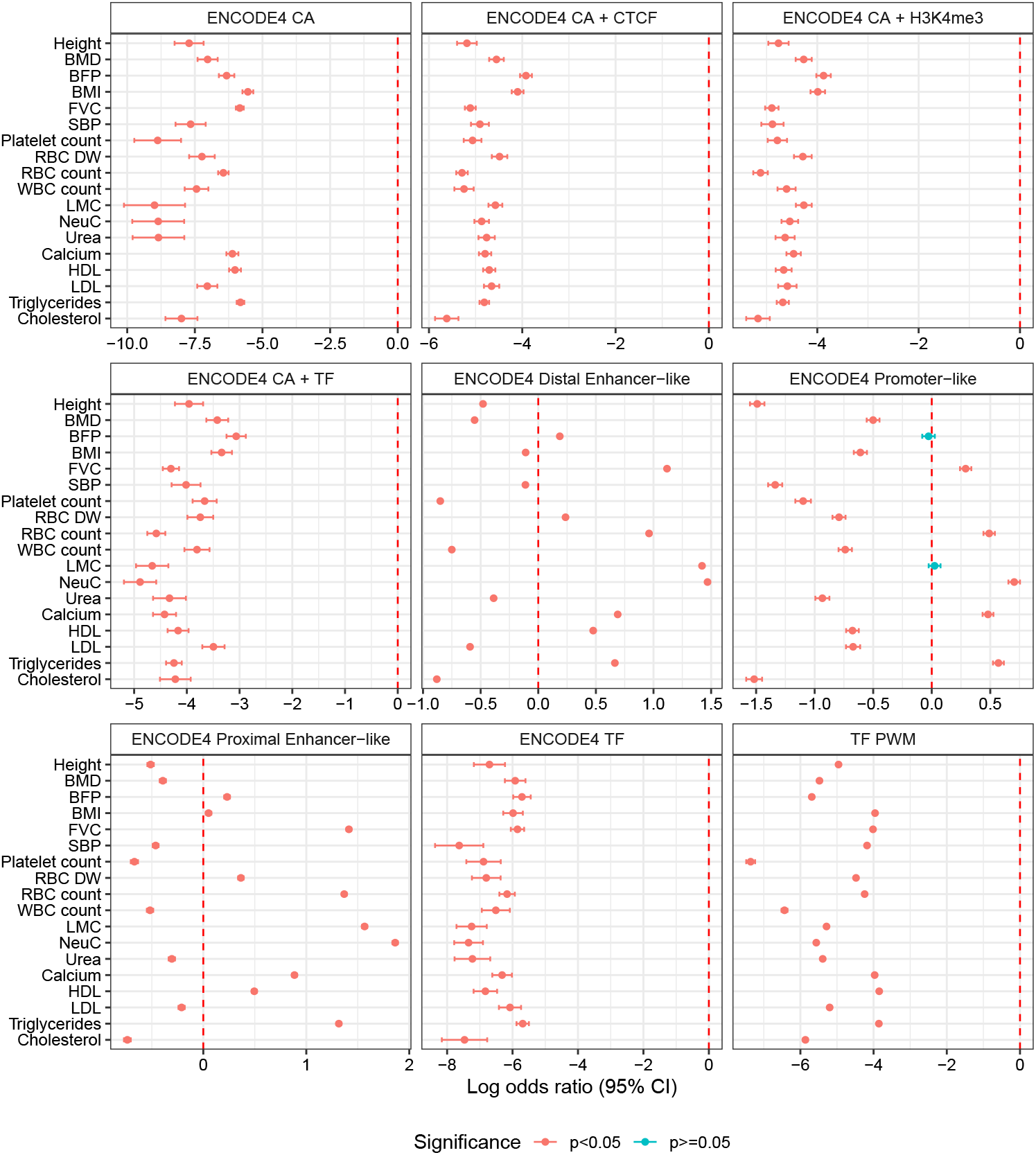
Logistic regression model coefficients – ENCODE4 and TF PWM

**Fig. C5.**
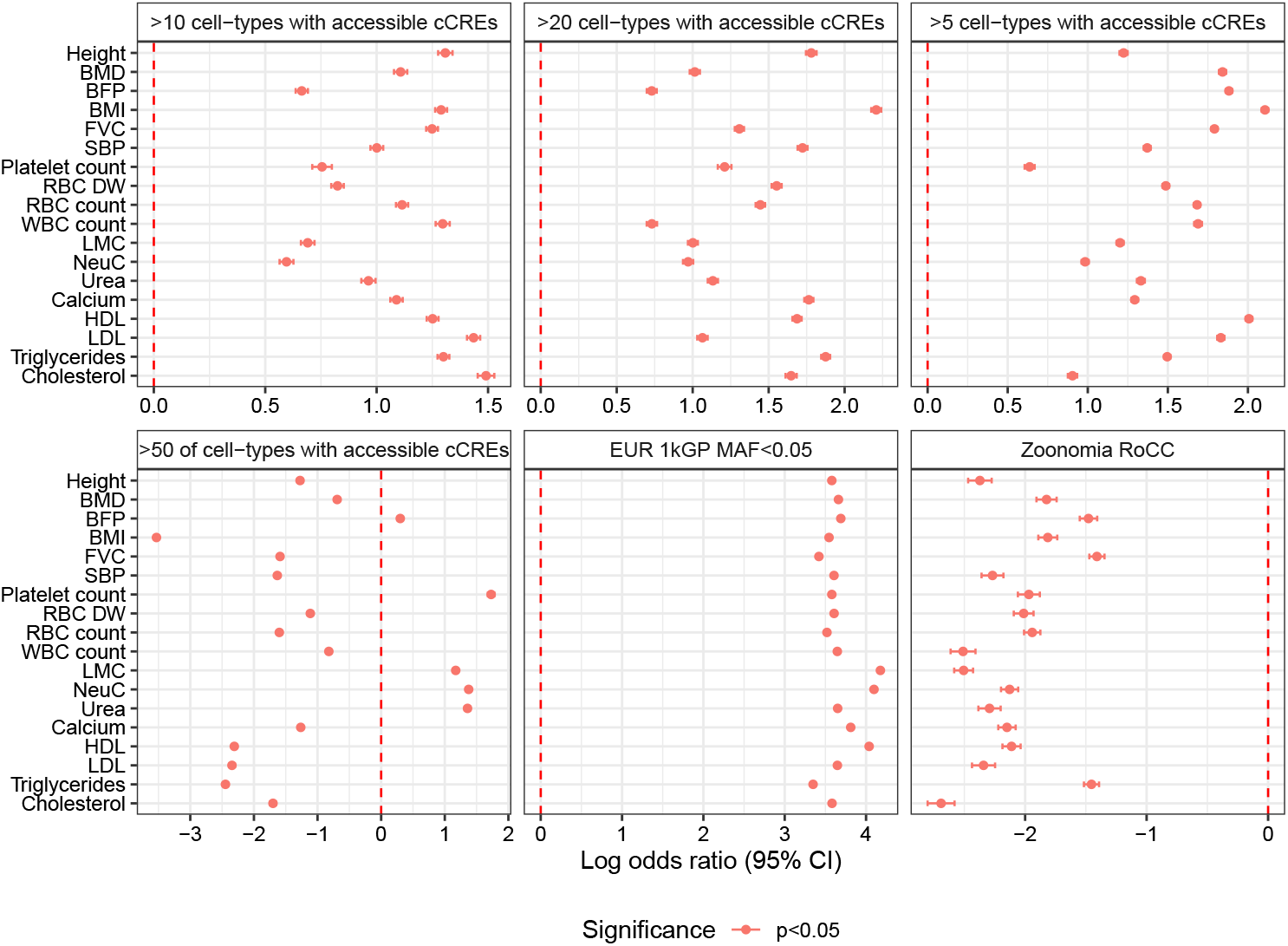
Logistic regression model coefficients – 1kGP, number of accessible cell types (cCRE) and Zoonomia

